# BOLD Amplitude Correlates of Preclinical Alzheimer’s Disease

**DOI:** 10.1101/2024.10.27.24316243

**Authors:** Stanislau Hrybouski, Sandhitsu R. Das, Long Xie, Christopher A. Brown, Melissa Flamporis, Jacqueline Lane, Ilya M. Nasrallah, John A. Detre, Paul A. Yushkevich, David A. Wolk

## Abstract

Alzheimer’s disease (AD) is characterized by a long preclinical stage during which molecular markers of amyloid beta and tau pathology rise, but there is minimal neurodegeneration or cognitive decline. Previous literature suggests that measures of brain function might be more sensitive to neuropathologic burden during the preclinical stage of AD than conventional measures of macrostructure, such as cortical thickness. However, among studies that used resting-state functional Magnetic Resonance Imaging (fMRI) acquisitions with Blood Oxygenation Level Dependent (BOLD) contrast, most employed connectivity-based analytic approaches, which discard information about the amplitude of spontaneous brain activity. Consequently, little is known about the effects of amyloid and tau pathology on BOLD amplitude. To address this knowledge gap, we characterized the effects of preclinical and prodromal AD on the amplitude of low-frequency fluctuations (ALFF) of the BOLD signal both at the whole-brain level and, at a more granular level, focused on subregions of the medial temporal lobe. We observed reduced ALFF in both preclinical and prodromal AD. In preclinical AD, amyloid positivity was associated with a spatially diffuse ALFF reduction in the frontal, medial parietal, and lateral temporal association cortices, while tau pathology was negatively associated with ALFF in the entorhinal cortex. These ALFF effects were observed in the absence of observable macrostructural changes in preclinical AD and remained after adjusting for structural atrophy in prodromal AD, indicating that ALFF offers additional sensitivity to early disease processes beyond what is provided by traditional structural imaging biomarkers of neurodegeneration. We conclude that ALFF may be a promising imaging-based biomarker for assessing the effects of amyloid-clearing immunotherapies in preclinical AD.

## INTRODUCTION

The field of Alzheimer’s disease (AD) is entering an exciting era of disease-modifying therapies. Recent FDA-approved immunotherapies target β-amyloid (Aβ) plaques, one of the hallmarks neuropathologies of AD, along with tau-containing neurofibrillary tangles. Clinical trials have shown that amyloid-clearing treatments can slow, but not halt, the rate of cognitive decline in patients with symptomatic disease (Sims et al., 2023; van Dyck et al., 2023). Since AD is characterized by a long preclinical stage, often lasting a decade or longer, during which molecular markers of AD rise, but there is minimal neurodegeneration and little, if any, cognitive decline (Jack et al., 2024), it is reasonable to expect that novel therapeutic treatments of AD, especially anti-amyloid immunotherapies, would be most effective in preclinical AD prior to the onset of amyloid-facilitated tau spread and subsequent neurodegeneration (Abramov et al., 2009; Hrybouski et al., 2023; Pooler et al., 2013; Schoonhoven et al., 2023; Sperling et al., 2023; Wu et al., 2016). Thus, there is a need for biomarkers sensitive to very early effects of disease pathology in the context of preclinical AD for disease staging and monitoring.

Amyloid plaques largely originate in brain regions that anatomically correspond to the default mode network (Buckner et al., 2005; Mormino et al., 2011; Palmqvist et al., 2017; Raichle et al., 2001). These typically include the medial frontal, orbitofrontal, posterior cingulate, and precuneus cortices. By the late preclinical stage, Aβ deposits are detectable in most of the neocortex (Cho et al., 2016; Palmqvist et al., 2017; Thal et al., 2002; Villeneuve et al., 2015). Because multiple studies of preclinical AD revealed the disruption of the default mode network in task-based and resting-state functional Magnetic Resonance Imaging (fMRI) experiments (Elman et al., 2016; Hedden et al., 2009; Mormino et al., 2011; Sheline et al., 2010; Sperling et al., 2009), it is thought that elevated Aβ has a deleterious effect on brain function fairly early in the course of the disease (Abramov et al., 2009; Berron et al., 2021; Hrybouski et al., 2023; Maass et al., 2019; Palop & Mucke, 2016).

Early cortical tau tangles, on the other hand, frequently emerge in the transentorhinal region of the medial temporal lobe (MTL), from which tau pathology spreads to other structures in the MTL, including the amygdala and hippocampus, and subsequently to the inferolateral temporal cortex (Braak et al., 2006; Braak & Braak, 1991; Therriault et al., 2022; Yassa, 2014; Yushkevich et al., 2021). As the disease progresses, tau spreads further throughout the temporal lobe and multiple neocortical structures (Braak et al., 2006; Therriault et al., 2022). Brain regions susceptible to tau accumulation also exhibit altered fMRI functional connectivity in early Alzheimer’s disease (Berron et al., 2020; Das et al., 2013; Elman et al., 2016; Hrybouski et al., 2023). As opposed to amyloid-associated changes in MTL function, which are frequently characterized by increased connectivity and activation (Adams et al., 2021; Das et al., 2013; Hrybouski et al., 2023; Stargardt et al., 2015), tau-related effects on MTL circuits are characterized by suppressed MTL function (Angulo et al., 2017; Busche et al., 2012).

Despite providing valuable insights into connectome-level changes in early AD, the utility of functional connectivity as a biomarker has been limited due to its low-to-moderate reproducibility at the individual level (Braun et al., 2012; Noble et al., 2019; Noble et al., 2017). Amplitude of Low-Frequency Fluctuations (ALFF) measures BOLD signal variability over time and is a complementary measure derived from resting-state BOLD fMRI data (Zang et al., 2007). Unlike functional connectivity, ALFF has excellent test-retest reliability and is stable over time (Braun et al., 2012; Cahart et al., 2023; X. Wang et al., 2013; Zuo et al., 2010). Because ALFF is derived from the BOLD signal, ALFF is related to fluctuations in cerebral blood flow (CBF), neurovascular coupling itself, and activity-dependent metabolic demands (Baller et al., 2022; Nugent et al., 2015; Wang et al., 2021). Previous studies documented ALFF changes in amnestic Mild Cognitive Impairment (MCI) and AD, as well as in individuals with subjective cognitive complaints (Gao et al., 2023; Han et al., 2011; Liu et al., 2014; Sun et al., 2016; Xi et al., 2013; Yang et al., 2018; Zhang et al., 2021; Zhao et al., 2014; Zhao et al., 2015; Zhou et al., 2015; Zhuang et al., 2012). However, these studies did not employ amyloid or tau molecular biomarkers of AD, and the effects of Aβ and tau pathology on ALFF are largely unexplored, especially in preclinical AD (Jack et al., 2024). We are aware of only one study (Millar et al., 2020) that examined the relationship between BOLD variability within spherical ROIs and CSF-based biomarkers in preclinical AD. Millar et al. (2020) reported a diffuse negative effect of amyloid on BOLD variability; however, they did not identify any statistically significant relationships between BOLD variability and pTau-181. The association between ALFF and tau-PET, which allows for regional quantification of tau burden in the MTL, remains unexplored in preclinical AD. The aim of our current study was to assess ALFF as a potential biomarker of functional abnormalities in preclinical and prodromal AD. To do so, we performed whole-brain voxelwise comparisons between amyloid-negative (Aβ−) cognitively unimpaired older adults and amyloid-positive (Aβ+) cognitively unimpaired individuals with preclinical AD or Aβ+ patients with MCI due to AD. We also examined the relationship between the MTL tau burden and ALFF in anatomically defined granular subregions of the MTL.

Previous studies revealed reduced glucose metabolism and cerebral blood flow (CBF) in AD (Alexopoulos et al., 2012; Alsop et al., 2000; Chételat et al., 2003; Friedland et al., 1989; Ishii et al., 1997; Minoshima et al., 2022; Mosconi et al., 2008; Silverman et al., 2001; Wolk & Detre, 2012), and amyloid-β is associated with CBF changes in cognitively unimpaired individuals (Mattsson et al., 2014; Sojkova et al., 2008). Since BOLD ALFF is positively associated with CBF and metabolic activity (Baller et al., 2022; Nugent et al., 2015; Wang et al., 2021), we predicted a negative relationship between cortical ALFF and amyloid deposition in a diffuse pattern mirroring that of amyloid deposition itself (Cho et al., 2016; Palmqvist et al., 2017; Villeneuve et al., 2015). On the other hand, since tau pathology in early AD is mainly confined to the MTL (Braak & Braak, 1991; Therriault et al., 2022), we reasoned that any functional changes driven by tau, especially during the preclinical stage of the disease, should be localized within MTL structures.

## MATERIALS AND METHODS

### Participants

In this cross-sectional study, we analyzed data from 204 individuals from the Aging Brain Cohort (ABC) study of the University of Pennsylvania Alzheimer’s Disease Research Center (Penn ADRC). Each of these participants undergoes annual cognitive evaluations, including psychometric testing as prescribed by the Uniform Data Set 3.0 (Weintraub et al., 2018). Consensus diagnoses are subsequently reached by an expert team of neurologists, neuropsychologists, and radiologists. Amyloid status was determined by a trained nuclear medicine physician based on visual reads of amyloid-PET scans using standard criteria. In this study we subdivided our participants into three groups: (i) Aβ-negative cognitively unimpaired (Aβ− CU) older adults, (ii) Aβ-positive cognitively unimpaired (Aβ+ CU) older adults with preclinical AD, and (iii) Aβ-positive older adults with MCI (Aβ+ MCI) due to AD (for detailed information about each group, see Table 1). The study was approved by the University of Pennsylvania Institutional Review Board.

**Table 1.** Demographic information for each group in the study.

| | Aβ- CU<br>(N = 131) | Aβ+ CU<br>(N = 33) | Aβ+ MCI<br>(N = 40) | Group Differences |
| --- | --- | --- | --- | --- |
| Age (years) |  |  |  |  |
| Mean ± SD | 71.0 ± 5.0 | 74.9 ± 6.0 | 73.3 ± 6.8 | Aβ- CU < Aβ+ CU *** |
| Range [min/max] | 59/87 | 65/87 | 57/87 | Aβ- CU < Aβ+ MCI * |
| Race |  |  |  |  |
| (White/Black/Asian/Mixed) | 85/44/0/2 | 27/6/0/0 | 36/4/0/0 | Aβ- CU different from Aβ+ MCI ** |
| Sex (males/females) | 46/85 | 10/23 | 18/22 | none |
| Education (years) ± SD | 16.3 ± 2.5 | 16.8 ± 1.8 | 17.0 ± 2.4 | none |
| Total MoCA ± SD | 27.0 ± 2.1 | 26.9 ± 2.4 | 21.3 ± 4.6 | Aβ+ MCI < others *** |
| CDR (sum/global) | 0.061/0.050 | 0.0937/0.078 | 2.150/0.525 | Aβ+ MCI > Aβ+ CU ***<br>Aβ+ MCI > Aβ- CU *** |
| Composite Florbetaben<br>SUVR ± SD (n) | 0.957 ± 0.088<br>(n = 120) | 1.297 ± 0.190<br>(n = 32) | 1.448 ± 0.126<br>(n = 39) | Aβ- CU < Aβ+ CU ***<br>Aβ- CU < Aβ+ MCI ***<br>Aβ+ CU < Aβ+ MCI *** |
| Transentorhinal Flortaucipir<br>SUVR ± SD (n) | 1.072 ± 0.101<br>(n = 99) | 1.209 ± 0.179<br>(n = 25) | 1.524 ± 0.199<br>(n = 11) | Aβ- CU < Aβ+ CU **<br>Aβ- CU < Aβ+ MCI ***<br>Aβ+ CU < Aβ+ MCI *** |

SUVR = standardized uptake value ratio, SD = standard deviation
\* $P < 0.05$ ; \*\* $P < 0.01$ ; \*\*\* $P < 0.001$
Independent samples $t$ -tests were used for group comparisons of the continuous variables, and $\chi^2$ -tests were used for comparisons of the categorical variables. Holm-Bonferroni corrected $p$ -values are reported.

### Image Acquisition

All structural and functional MRI scans were acquired using a 3.0 T Siemens Prisma scanner (Erlangen, Germany) at the Center for Advanced Magnetic Resonance Imaging and Spectroscopy (University of Pennsylvania, Philadelphia, PA). During functional scans, participants underwent an eyes-open T2*-sensitive Gradient Echo Planar Imaging (EPI) resting-state acquisition [repetition time (TR): 720 ms; echo time (TE): 37 ms; flip angle: 52°; field of view: 208 × 208 mm^2^; voxel size: 2 × 2 × 2 mm^3^; 72 interleaved slices; phase encoding direction: anterior to posterior; partial k-space: 7/8; multi-band acceleration factor: 8], during which they passively viewed a static fixation cross. For structural analyses, a whole-brain T1-weighted structural scan was acquired using a 3D Magnetization Prepared Rapid Gradient Echo sequence [TR: 2400 ms; TE: 2.24 ms; inversion time: 1060 ms; flip angle: 8°; field of view: 256 × 240 × 167 mm^3^; voxel size: 0.8 × 0.8 × 0.8 mm^3^]. To estimate B0 inhomogeneity, either gradient echo [TR: 580 ms; TE1/TE2: 4.12/ 6.58 ms; flip angle: 45°; field of view: 240 × 240 mm2; voxel size: 3 × 3 × 3 mm^3^; 60 interleaved slices] or spin echo field maps [TR: 8000 ms; TE: 66 ms; flip angle: 90°; refocus flip angle: 180°; field of view: 208 × 208 mm^2^; voxel size: 2 × 2 × 2 mm^3^; 72 interleaved slices; phase encoding directions: anterior-to-posterior and posterior-to-anterior] were also acquired. Gradient echo field maps were used in 196 out of 204 participants.

During amyloid-PET scans, participants underwent a 20-minute brain scan (four frames of 5-min duration) following an injection of 8.1 mCi ± 20% of ^18^F-florbetaben or 10 mCi ± 20% of ^18^F-florbetapir and a standard uptake phase (90 min for ^18^F-florbetaben and 50 min for ^18^F-florbetapir). Tau-PET was obtained using six 5-minute frames from 75 to 105 min after the injection of 10 mCi ± 20% of ^18^F-flortaucipir.

### Functional image preprocessing

The initial preprocessing of functional data was performed on the Flywheel Imaging Informatics Platform (http://flywheel.io) using the fMRIprep 20.0.7 pipeline (Esteban et al., 2019; Gorgolewski et al., 2011). A detailed description of this preprocessing pipeline has been provided elsewhere (Esteban et al., 2019; Hrybouski et al., 2023). Briefly, we performed rigid-body realignment and field-map-based distortion correction (Cox & Hyde, 1997; Jenkinson et al., 2002; Jezzard & Balaban, 1995) followed by functional-to-structural rigid-body boundary-based registration (Greve & Fischl, 2009) and SyN diffeomorphic registration to the 2-mm MNI152 template using Advanced Normalization Tools (ANTS; Avants & Gee, 2004; Avants et al., 2008). Warped fMRI datasets were smoothed with a 6-mm FWHM Gaussian Kernel and a linear decomposition of the data using Probabilistic Spatial Independent Component Analysis (ICA) was performed. Next, manual ICA-based denoising was performed by a single rater (SH) with the aid of the automated movement component classifier, ICA-AROMA (Pruim et al., 2015). To ensure T1 equilibrium, the first 10 fMRI volumes from each resting-state scan were discarded, and 32 CompCorr nuisance regressors (Behzadi et al., 2007) were generated from White Matter (WM) and Cerebrospinal (CSF) signals using eroded WM/CSF masks: 16 regressors were extracted from WM and 16 from CSF. CompCorr regressors were supplemented with a 24P Friston motion set (Friston et al., 1996) and six low-frequency sine and cosine waves (periods of π, 2π, and 3π over 410 time points). Time points impacted by excessive head motion or sudden signal spikes (standardized DVARS-score > 1.5; Power et al., 2014) were removed from the data using the spike regression approach (Lemieux et al., 2007; Satterthwaite et al., 2013). Finally, the fMRI datasets were low-pass filtered to remove frequencies above 0.15 Hz, and any residual linear drifts were regressed out of the data. The preprocessed fMRI images were rescaled to percent signal change units (i.e., each participant’s average fMRI signal across all voxels within the brain and across all time points = 100).

Voxel-level ALFF was estimated using default settings in the Conn Toolbox (v. 22.a; Whitfield-Gabrieli & Nieto-Castanon, 2012). To compute ALFF, the filtered time course for each voxel is initially transformed into the frequency domain using a Fast Fourier Transform (Zang et al., 2007). The square root of the power spectrum is then calculated and averaged across the chosen frequency band (0.003-0.15 Hz, in the current study). Here, we computed raw, as opposed to normalized, ALFF because raw ALFF does not remove global amplitude differences that might be driven by diffuse neuropathologic changes, including Aβ. Mean ALFF map, representing raw ALFF values, averaged across participants is available in the Supplementary Materials (Suppl. Fig. 1)

### Structural image processing

The MTL subregions were segmented using the automated segmentation of hippocampal subfields-T1 (ASHS-T1) pipeline (Xie et al., 2019). ASHS-T1 segments the MTL into Brodmann Area 36 (BA36), Brodmann Area 35 (BA35), entorhinal cortex (ERC), parahippocampal cortex (PHC), anterior hippocampus (aHP), and posterior hippocampus (pHP). Each subject’s ASHS-T1 MTL labels were registered to MNI space using the same deformation fields that were used to register that subject’s fMRI data to the 2-mm MNI152 template. An example ASHS-T1 segmentation is provided in the Supplementary Materials (Suppl. Fig. 2). The anatomical MRI was also parcellated into cortical, subcortical, and cerebellar ROIs using a multi-atlas segmentation method that is detailed elsewhere (Asman & Landman, 2013; Klein & Tourville, 2012; Landman & Warfield, 2012; H. Wang et al., 2013; Wang & Yushkevich, 2013). Next, brain-wide cortical thickness maps were estimated using the ANTs cortical thickness pipeline, which uses a diffeomorphic registration-based thickness estimation method, DiReCT (Das et al., 2009). These thickness maps were subsequently registered to the 1-mm MNI152 template using the same deformation fields that we used to register fMRI images to MNI space. Prior to performing group comparisons of cortical thickness, we smoothed warped thickness maps with a 3-mm FWHM Gaussian Kernel. Since we used cortical thickness maps as a voxelwise covariate in some statistical analyses of ALFF, we generated a separate set of thickness maps that were registered to the 2-mm MNI152 template and were smoothed with a 6-mm FWHM kernel to ensure spatial consistency with the fMRI data. Thickness maps from six participants (five from the Aβ− CU group and one from the Aβ+ CU group) did not pass quality control and were excluded from statistical analyses. The intracranial volume (ICV) was computed using an in-house ICV segmentation software implemented using the ASHS codebase (Xie et al., 2019).

### PET image analysis

PET data were processed with in-house software. First, attenuation-corrected dynamic image frames were motion-corrected using *mcflirt* rigid-body registration (FSL 5.0.9; Jenkinson et al., 2002; Smith et al., 2004). The resulting motion-corrected PET frames were averaged and aligned with participants’ T1-weighted structural MRI scans using ANTs rigid-body registration with a mutual information metric (Avants & Gee, 2004; Avants et al., 2008). For tau PET, mean tracer uptake in the inferior cerebellar gray matter was computed and used as a reference to generate a standardized uptake value ratio (SUVR) map for the entire brain. Because our sample consisted primarily of CU individuals, tau-PET SUVR was extracted from the extrahippocampal anterior MTL region using ERC + BA35 combined ROIs from the previously described ASHS-T1 segmentations. For amyloid PET, mean tracer uptake in the whole cerebellum was computed and used as a reference to generate an SUVR map for the entire brain. A composite ROI consisting of the middle frontal, anterior cingulate, posterior cingulate, inferior parietal, precuneus, supramarginal, middle temporal, and superior temporal cortical regions was used to compute the global amyloid burden in each participant (Landau et al., 2013).

### Statistical analyses of ALFF and cortical thickness

General Linear Models (GLMs) were used to compare voxelwise ALFF among the three groups (i.e., Aβ− CU, Aβ+ CU, and Aβ+ MCI). In all voxelwise analyses, age, sex, as well as four variables summarizing in-scanner head motion during the fMRI scan – mean filtered framewise displacement (FD), standard deviation (SD) of filtered FD, maximum standardized DVARS, and SD of standardized DVARS – were used as covariates (Power et al., 2014). Filtered FD was generated from filtered realignment parameters. Realignment parameter filtering was performed using MATLAB scripts provided by Gratton et al. (2020). This step is recommended for fMRI acquisitions with sub-second temporal resolution because raw realignment parameters in such datasets are contaminated by breathing-associated magnetic field perturbations (Fair et al., 2020; Gratton et al., 2020), and raw FD metrics do not provide an accurate measure of head movement during the fMRI scan. In select voxelwise GLMs, cortical thickness was used as an additional voxel-specific covariate. The threshold-free cluster enhancement (TFCE) method from *randomise* function in FSL 6.0.4 was used for statistical inference (Smith & Nichols, 2009). In each test, the distribution of the TFCE statistic under the null hypothesis was estimated using 25,000 Freedman and Lane (1983) permutations. Statistical significance was determined using one-sided tests with α = 0.025.

To analyze the relationship between ALFF and AD pathology within the MTL, we computed mean ALFF scores for each of the ASHS-T1 MTL ROIs and averaged them across hemispheres. ROI-level GLMs were used to test for statistical associations between ALFF and MTL tau in each MTL ROI. To ensure that the time lag between the fMRI and tau PET scans did not confound our results, we incorporated time between them as a nuisance covariate and excluded individuals with > 5-year intervals between the two scans [one participant excluded]. We also performed a supplementary set of analyses on subjects with < 1.5-year intervals between tau-PET and MRI [122/136 (∼90%) of all participants with a tau PET]. In addition to the time lag between the fMRI and tau PET scans (subsequently PET-MRI Δ*t*), we also included age, sex, and the four head movement summary metrics as nuisance covariates. Structural measures (thickness for BA36, BA35, ERC, and PHC; volume + ICV for aHP and pHP) and amyloid status were also used as additional covariates in some analyses. Analyses that adjusted for amyloid status also adjusted for amyloid status × PET-MRI Δ*t* interaction. Statistical significance was assessed using 25,000 permutation tests. Bonferroni correction for multiple hypothesis testing (6 tests) was used to control the family-wise error (FWE) rate. These intra-MTL GLM analyses were performed using custom MATLAB scripts. Group comparisons of intra-MTL ALFF were performed similarly. Where appropriate, *post hoc* mediation analyses were performed using the Mediation Toolbox for MATLAB (Wager et al., 2008). In those mediation analyses, we used the non-parametric bootstrap with 100,000 samples to construct bias-corrected accelerated (BCa) confidence intervals for statistical tests on direct and indirect effects (Kenny et al., 2003; Shrout & Bolger, 2002).

## RESULTS

### ALFF is reduced in early Alzheimer’s disease

Relative to the Aβ− CU group, individuals with preclinical AD displayed significantly lower ALFF in multiple cortical regions: medial and inferior frontal, orbitofrontal, posterior cingulate, precuneus, insula, primary somatomotor, supplementary motor, medial visual, lateral temporal, medial temporal, and temporal pole (Fig. 1a). In addition, the preclinical (i.e., Aβ+ CU) group had lower ALFF in the bilateral striatum, thalamus, amygdala, and anterior hippocampus (Fig. 1a, slice view). Across all voxels that showed a statistically significant ALFF difference between the Aβ+ CU and Aβ− CU groups, ALFF in the Aβ+ CU group was ≈12% lower than in the Aβ− CU group (*M*_Aβ− CU_ = 0.215, *M*_Aβ+ CU_ = 0.189).

**Figure 1.**
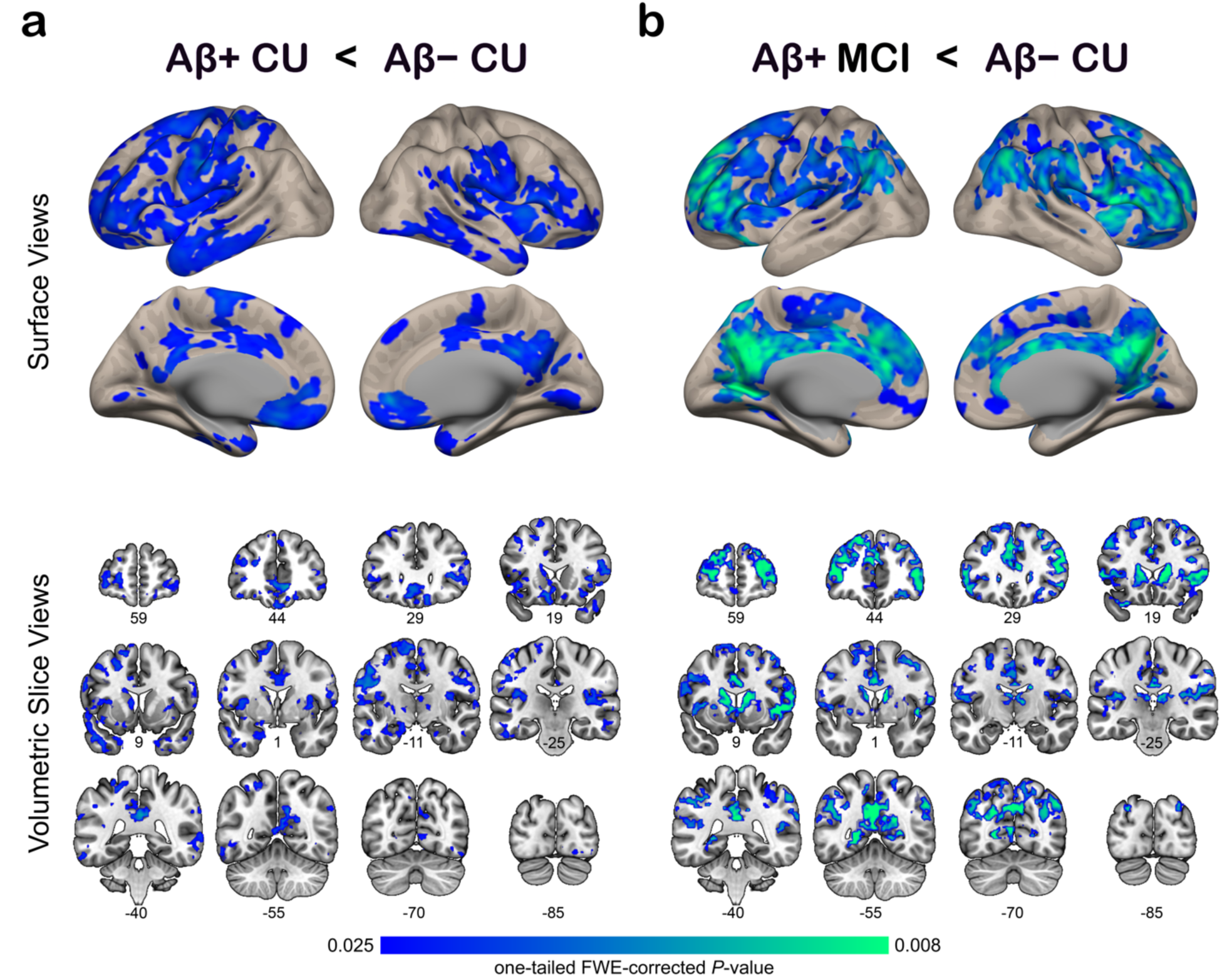
Voxelwise ALFF differences between preclinical (a) or prodromal (b) AD groups and age-matched controls. Group comparisons were performed using voxelwise GLMs with age, sex, and head movement metrics as nuisance covariates. The color map corresponds to FWE-corrected TFCE *P-*values. Slice coordinates in volumetric panels correspond to MNI152 space. Abbreviations: AD = Alzheimer’s disease; ALFF = amplitude of low-frequency fluctuations; CU cognitively unimpaired; MCI = mild cognitive impairment; TFCE = threshold-free cluster enhancement.

Next, we compared ALFF of the Aβ+ MCI and Aβ− CU groups. The Aβ+ MCI group displayed reduced ALFF in most of the frontal association cortex with the inferior medial region, including orbitofrontal, as the notable exception with preserved ALFF (Fig. 1b, surface view). Reduced ALFF in the Aβ+ MCI group was also observed in the thalamus and striatum, and in the bilateral somatomotor, supplementary motor, insular, posterior cingulate, precuneus, and lateral parietal cortices (Fig. 1b). Across all voxels that showed a statistically significant ALFF difference between the Aβ+ MCI and Aβ− CU groups, the Aβ+ MCI group displayed ≈13% lower ALFF (*M*_Aβ− CU_ = 0.226, *M*_Aβ+ MCI_ = 0.200). No statistically significant differences were observed when comparing the prodromal and preclinical AD groups with each other.

To test whether group-associated differences in ALFF could be explained by AD-associated structural atrophy, we (1) compared cortical thickness maps of the three groups and (2) repeated ALFF group comparisons on thickness-adjusted ALFF maps. Consistent with previously published literature, we observed reduced cortical thickness in the Aβ+ MCI group relative to the other two groups in typical AD regions, including limbic and medial parietal cortices (Fig. 2). Importantly, no thickness differences were observed between Aβ− CU and Aβ+ CU groups, and repeating ALFF group comparisons on thickness-adjusted ALFF maps did not eliminate the effect of reduced ALFF in the Aβ+ CU group. Together, these results indicate that in preclinical AD, the functional measure of ALFF might be more sensitive to AD-related pathology than cortical thickness.

**Figure 2.**
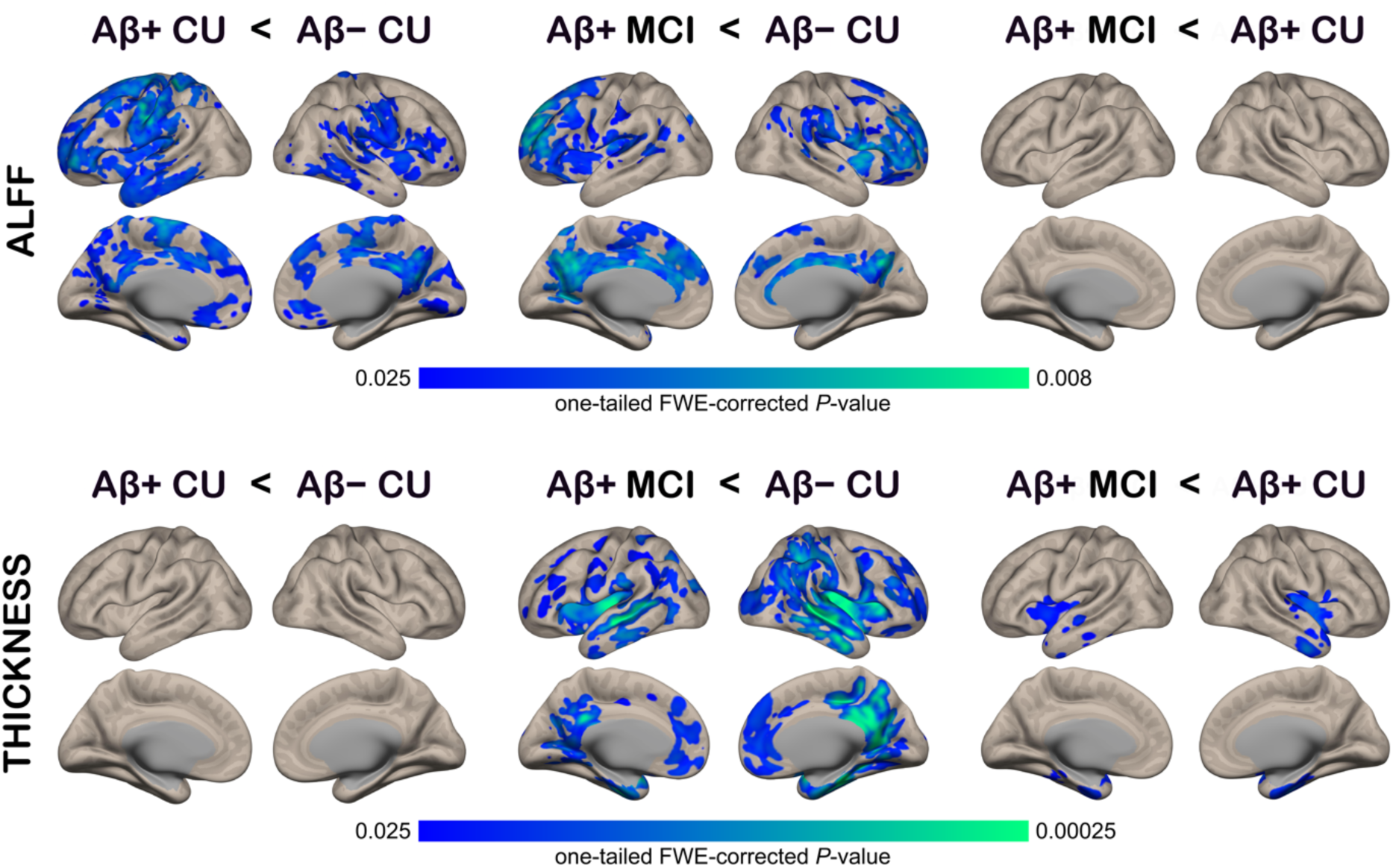
Matched pairwise group comparisons of thickness-adjusted ALFF (top) and cortical thickness (bottom). Voxelwise GLMs were used for statistical testing with age, sex, and head movement metrics as nuisance covariates in all GLMs. Local cortical thickness was included as an additional voxel-specific covariate in the ALFF comparisons. Only TFCE FWE-corrected results are shown. Abbreviations: AD = Alzheimer’s disease; ALFF = amplitude of low-frequency fluctuations; CU = cognitively unimpaired; MCI = mild cognitive impairment; TFCE = threshold-free cluster enhancement.

### Entorhinal ALFF is Negatively Associated with MTL Tau

Examining the relationship between MTL tau burden and subregional MTL ALFF for each of the ASHS-T1 ROIs revealed a statistically significant negative association between MTL tau mean SUVR and ERC ALFF (*β* = −0.224, corrected *P* = 0.013; Table 2, Fig. 3a). Statistical adjustment for structural variation strengthened this effect (*β* = −0.281, corrected *P* < 0.001; Table 2; Fig. 3a), while adjustment for amyloid positivity did not eliminate it (Table 2, Fig. 3a). No statistically significant associations between ALFF and MTL tau SUVR were observed in the other MTL ROIs, although there was universally a negative relationship across regions, with some reaching trend-level significance (Table 2).

**Figure 3.**
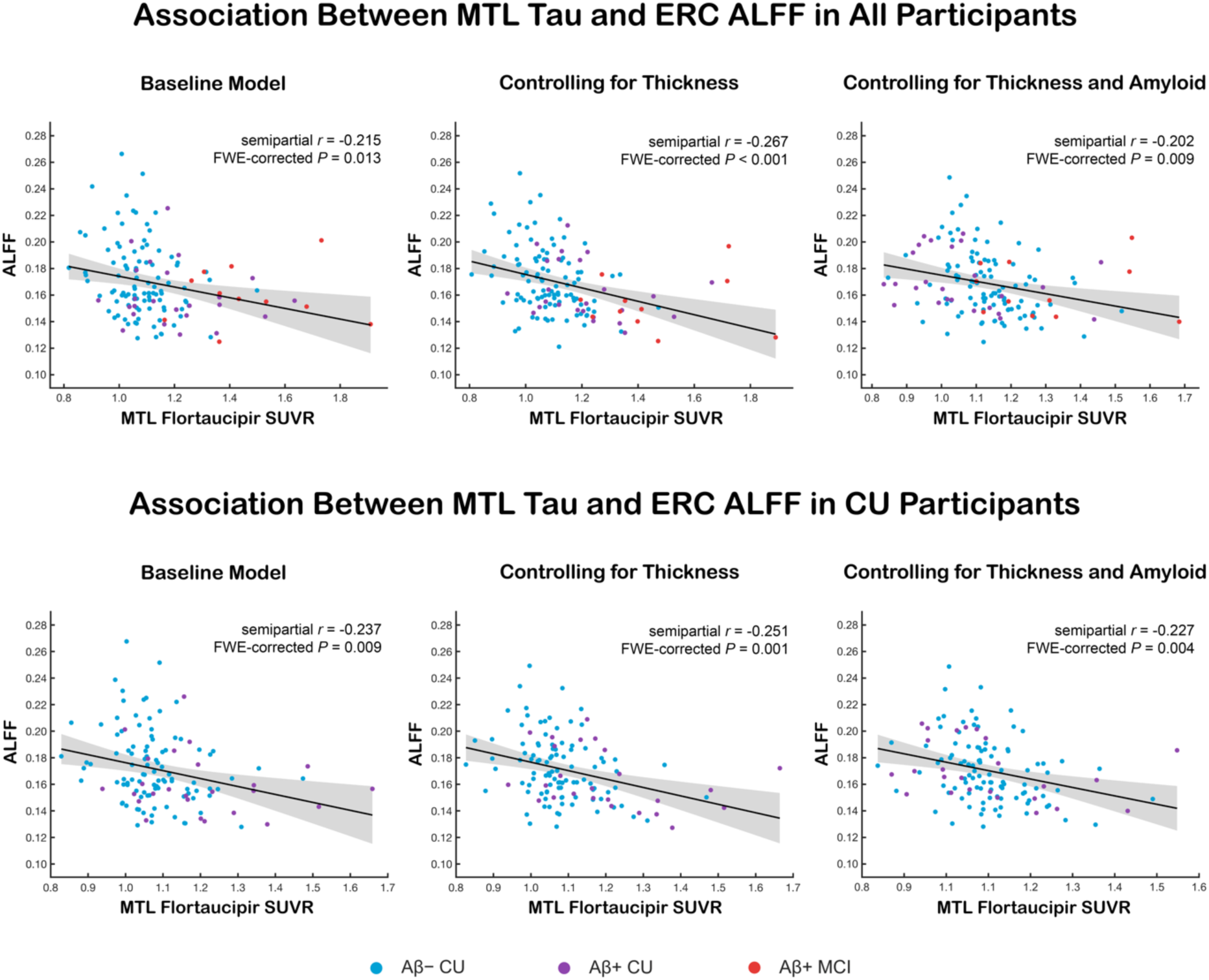
Added value plots depicting the relationship between MTL tau and ERC ALFF in all participants with tau PET (top panels) and in those who were cognitively unimpaired (CU) at the time of the scan (bottom panels). All baseline models were adjusted for age, sex, head movement, and time interval between the fMRI and tau-PET scans (MRI-PET Δ*t*). The other models also adjusted for ERC thickness. The ‘Thickness + Amyloid’ model additionally adjusted for amyloid positivity and amyloid × MRI-PET Δ*t* interaction. Shaded areas around each fit correspond to 95% confidence intervals of the estimate. Abbreviations: AD = Alzheimer’s disease, ALFF = amplitude of low-frequency fluctuations, ERC = entorhinal cortex, MTL = medial temporal lobe.

**Table 2.** Association between transentorhinal tau and MTL ALFF.

|  | BA36 | BA35 | ERC | aHP | pHP | PHC |
| --- | --- | --- | --- | --- | --- | --- |
| **Baseline model ($N = 135$, $df = 126$)** |  |  |  |  |  |  |
| standardized $\beta$ | -0.128 | -0.154 | -0.224 | -0.155 | -0.133 | -0.137 |
| $t$-statistic | -1.611 | -1.933 | -2.975 | -1.986 | -1.705 | -1.787 |
| FWE-corrected $P$ | 0.266 | 0.150 | **0.013** | 0.133 | 0.226 | 0.195 |
| **Controlling for structural measures ($N = 135$, $df = 124/125$)** |  |  |  |  |  |  |
| standardized $\beta$ | -0.135 | -0.190 | -0.281 | -0.150 | -0.171 | -0.145 |
| $t$-statistic | -1.690 | -2.433 | -4.203 | -2.264 | -2.405 | -1.943 |
| FWE-corrected $P$ | 0.280 | 0.062 | **< 0.001** | 0.090 | 0.065 | 0.177 |
| **Controlling for structural measures and amyloid status ($N = 135$, $df = 122/123$)** |  |  |  |  |  |  |
| standardized $\beta$ | -0.031 | -0.132 | -0.259 | -0.131 | -0.149 | -0.066 |
| $t$-statistic | -0.319 | -1.388 | -3.185 | -1.621 | -1.745 | -0.727 |
| FWE-corrected $P$ | 0.998 | 0.456 | **0.009** | 0.316 | 0.256 | 0.902 |

Abbreviations: ALFF = amplitude of low-frequency fluctuations, aHP = anterior hippocampus, BA36/35 = Brodmann area 36/35, ERC = entorhinal cortex, MTL = medial temporal lobe, PHC = parahippocampal cortex, pHP = posterior hippocampus.

To test whether intra-MTL ALFF was associated with tau burden in the MTL when limited to CU participants, we repeated tau-ALFF relationship analyses in CU participants. As above, we observed a negative relationship between MTL tau and ALFF in the ERC (*β* = −0.242, corrected *p* = 0.009; Table 3; Fig. 3b) that was strengthened after controlling for inter-individual differences in ERC thickness (*β* = −0.256, corrected *p* = 0.001, Table 3, Figure 3b). Importantly, the effect of tau on rs-fMRI ALFF was detectable in the absence of tau-structure associations in CU older adults and in the absence of MTL structural differences between control and preclinical groups (all *p*s > 0.1, Tables 1-2). Controlling for amyloid positivity using amyloid status as a nuisance covariate confirmed that in CU older adults, ERC ALFF was associated with tau tracer uptake in the MTL independent of amyloid status (Table 3, Fig. 3b). Restricting the analyses to participants with < 1.5-year gap between MRI and tau PET scans did not alter these results (Suppl. Tables 3-4). Voxelwise tests for association between MTL tau and ALFF also revealed a trend-level effect (one-tailed α = 0.050) in left ERC (Suppl. Fig. 3).

**Table 3.** Association between transentorhinal tau and MTL ALFF in cognitively unimpaired older adults.

|  | BA36 | BA35 | ERC | aHP | pHP | PHC |
| --- | --- | --- | --- | --- | --- | --- |
| **All CU: Baseline model ($N = 124$, $df = 115$)** |  |  |  |  |  |  |
| standardized $\beta$ | -0.150 | -0.164 | -0.242 | -0.177 | -0.121 | -0.126 |
| $t$-statistic | -1.819 | -1.997 | -3.138 | -2.210 | -1.489 | -1.579 |
| FWE-corrected $P$ | 0.185 | 0.131 | 0.009 | 0.084 | 0.326 | 0.282 |
| **All CU: Controlling for structural measures ($N = 124$, $df = 113/114$)** |  |  |  |  |  |  |
| standardized $\beta$ | -0.160 | -0.176 | -0.256 | -0.147 | -0.135 | -0.138 |
| $t$-statistic | -1.940 | -2.201 | -3.837 | -2.176 | -1.933 | -1.797 |
| FWE-corrected $P$ | 0.179 | 0.105 | 0.001 | 0.109 | 0.178 | 0.233 |
| **All CU: Controlling for structural measures and amyloid status ($N = 124$, $df = 111/112$)** |  |  |  |  |  |  |
| standardized $\beta$ | -0.107 | -0.148 | -0.256 | -0.146 | -0.136 | -0.101 |
| $t$-statistic | -1.184 | -1.685 | -3.468 | -1.971 | -1.769 | -1.196 |
| FWE-corrected $P$ | 0.589 | 0.284 | 0.004 | 0.166 | 0.241 | 0.581 |

Abbreviations: ALFF = amplitude of low-frequency fluctuations, aHP = anterior hippocampus, BA36/35 = Brodmann area 36/35, ERC = entorhinal cortex, MTL = medial temporal lobe, PHC = parahippocampal cortex, pHP = posterior hippocampus.

Taken together, our results indicate a robust association between tau accumulation in the transentorhinal cortex and ERC ALFF.

### Differential Effects of Amyloid and Tau on Entorhinal vs. Medial Parietal ALFF

To further determine whether AD-associated effects on ALFF in the MTL were driven primarily by tau, while those in the neocortical regions were driven primarily by amyloid, we examined the relationship between these two pathologies and ALFF in brain areas that are known to be more vulnerable to amyloid than tau and vice versa. Previous amyloid PET literature has revealed that the medial parietal cortex is especially vulnerable to amyloid accumulation, whereas tau pathology originates in the transentorhinal region of the MTL. Thus, we focused on those two brain areas when evaluating amyloid vs. tau contributions to AD-associated changes in ALFF.

We used statistical maps from our previous ALFF group comparisons (i.e., Aβ− CU vs. Aβ+ CU and Aβ− CU vs. Aβ+ MCI with and without voxelwise thickness covariates; Figs. 1-2) to select medial parietal regions with AD-associated ALFF effects. First, we identified clusters with statistically significant ALFF group effects either in the precuneus or posterior cingulate. The resulting cluster maps were binarized and used to create a set of intersection clusters representing medial parietal regions with consistent AD-associated ALFF effects in preclinical and symptomatic AD (see Suppl. Fig. 4 for ROI visuals). We chose ERC as the tau-vulnerable ROI because tau pathology in this area emerges early in the disease, our above analyses supported a relationship with MTL tau tracer uptake, and it was the only MTL region to show amyloid-based group differences in ALFF (Table 4, Fig. 4).

**Figure 4.**
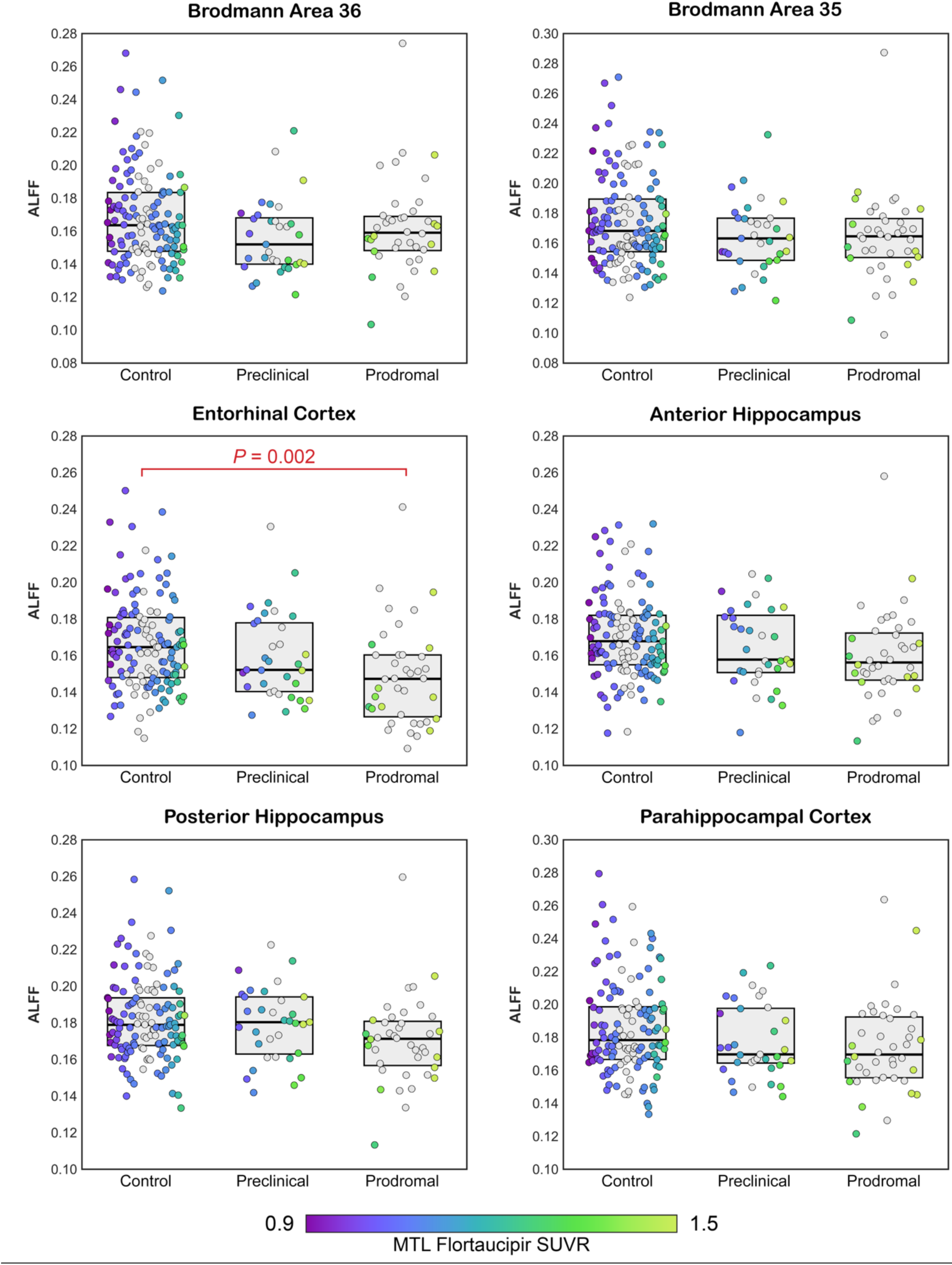
ALFF group differences in each MTL subregion. Box plots and individual data points represent ROI-averaged ALFF values after adjusting for age, sex, head movement, and structural measurements (volume + ICV for the hippocampal ROIs, thickness for others). Color of each data point represents that participant’s tau burden in the MTL. Data points from individuals who underwent amyloid-PET but not tau-PET scans are depicted in gray. Statistically significant pairwise comparisons are highlighted. Abbreviations: ALFF = amplitude of low-frequency fluctuations, MTL = medial temporal lobe.

**Table 4.** Differences in MTL ALFF among groups.

|  | BA36 | BA35 | ERC | aHP | pHP | PHC |
| --- | --- | --- | --- | --- | --- | --- |
| **Baseline model ($N = 204$, $df_1 = 2$, $df_2 = 195$)** |  |  |  |  |  |  |
| $M_{\text{control}}$ ($N = 131$) | 0.167 | 0.173 | 0.164 | 0.168 | 0.180 | 0.184 |
| $M_{\text{preclinical}}$ ($N = 33$) | 0.155 | 0.162 | 0.155 | 0.156 | 0.171 | 0.178 |
| $M_{\text{prodromal}}$ ($N = 40$) | 0.165 | 0.169 | 0.157 | 0.161 | 0.170 | 0.178 |
| $F$-statistic | 2.194 | 1.888 | 1.752 | 2.822 | 2.399 | 1.095 |
| FWE-corrected $P$ | 0.279 | 0.356 | 0.395 | 0.167 | 0.235 | 0.639 |
| **Baseline model + structural covariates ($N = 204$, $df_1 = 2$, $df_2 = 193/194$)** |  |  |  |  |  |  |
| $M_{\text{control}}$ ($N = 131$) | 0.168 | 0.174 | 0.166 | 0.170 | 0.182 | 0.186 |
| $M_{\text{preclinical}}$ ($N = 33$) | 0.156 | 0.164 | 0.159 | 0.164 | 0.179 | 0.178 |
| $M_{\text{prodromal}}$ ($N = 40$) | 0.162 | 0.163 | 0.149 | 0.160 | 0.170 | 0.174 |
| $F$-statistic | 2.218 | 2.624 | 5.878 | 2.452 | 2.606 | 2.534 |
| FWE-corrected $P$ | 0.333 | 0.242 | 0.015 | 0.274 | 0.242 | 0.259 |

Baseline models included age, sex, and head movement measures as covariates. Abbreviations: ALFF = amplitude of low-frequency fluctuations, aHP = anterior hippocampus, BA36/35 = Brodmann area 36/35, ERC = entorhinal cortex, MTL = medial temporal lobe, PHC = parahippocampal cortex, pHP = posterior hippocampus.

Next, we performed causal mediation analyses to test whether amyloid-associated effects on medial parietal and entorhinal ALFF were driven by local tau burden. Age, sex, regional cortical thickness, head movement summary metrics, PET-MRI Δ*t*, and amyloid status × PET-MRI Δ*t* interaction were used as covariates. We used a non-parametric bootstrap with 100,000 samples to construct bias-corrected accelerated (BCa) confidence intervals for statistical tests on direct and indirect effects. Mediation results showed that 86% of the effect of amyloid on ERC ALFF was mediated by MTL tau (Fig. 5a, top panel). Such mediation was not present in the medial parietal cortex (Fig. 5b, top panel). In the alternative model, ALFF was a marginal mediator (< 10%) of the effect of amyloid on MTL but not parietal tau (Fig. 5, bottom panels). Together, these results highlight the dissociable effects of amyloid and tau on resting-state ALFF.

**Figure 5.**
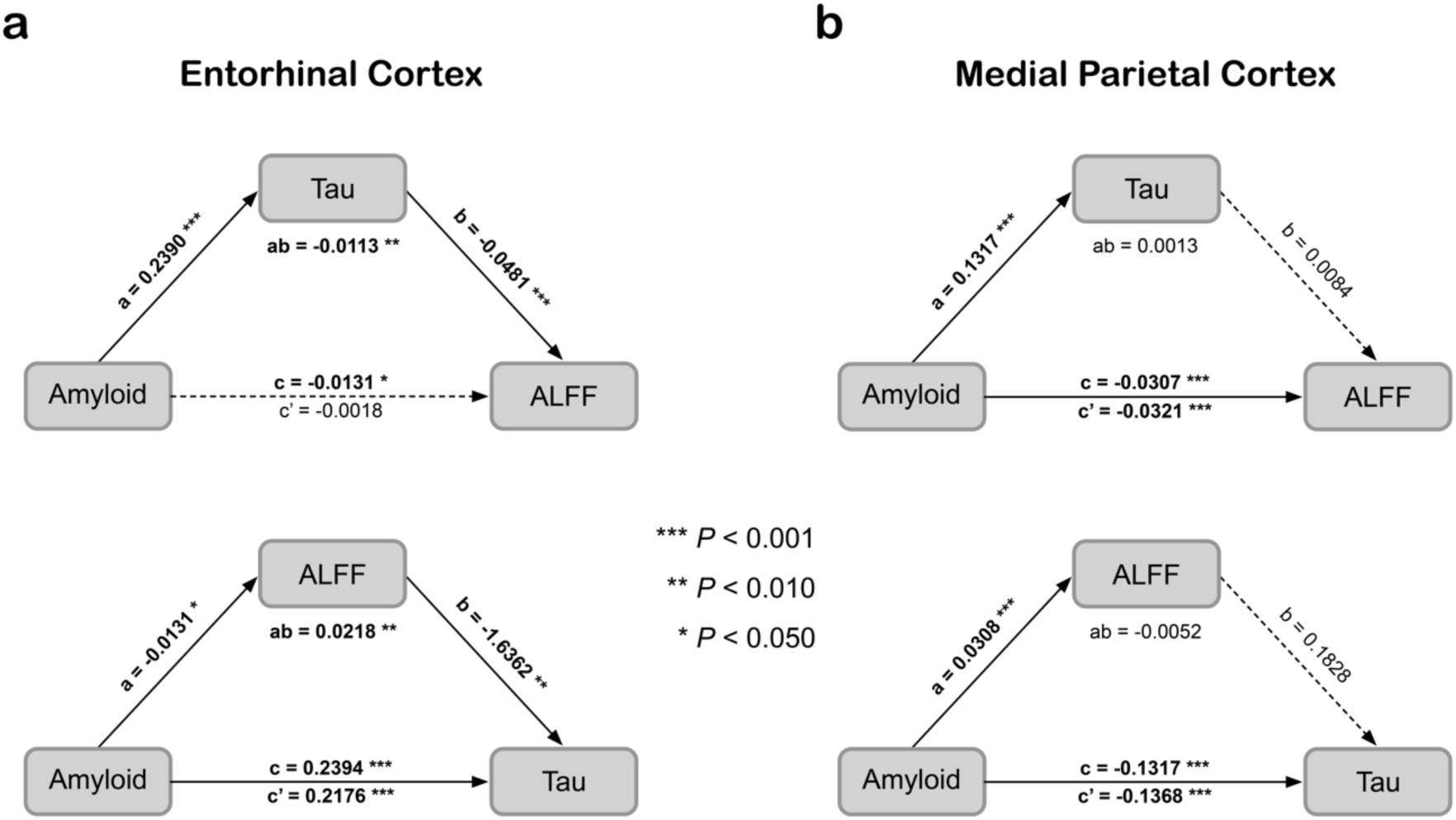
Mediation analyses for models investigating the relationships among amyloid status (i.e., Aβ- or Aβ-negative), regional tau burden, and entorhinal (**a**) versus medial parietal (**b**) ALFF. In the entorhinal cortex (ERC), the effect of amyloid on ALFF was fully mediated via MTL tau. In the alternative model, the effect of amyloid on MTL tau was marginally mediated by ERC ALFF. There were no such mediations in the medial parietal cortex. In all models, both the mediator and the outcome models were adjusted for age, sex, head movement, cortical thickness, and time lag between the fMRI and tau-PET scans. Numbers above or below the mediation path arrows represent linear regression coefficients for the corresponding associations. Abbreviations: ALFF = amplitude of low-frequency fluctuations; MTL = medial temporal lobe.

## DISCUSSION

This study evaluated the effects of both amyloid and tau measured by PET imaging on fMRI-based cortical and MTL ALFF in early AD. The following noteworthy findings emerged in this study: (1) AD pathology was associated with reduced resting-state fMRI ALFF in both preclinical and prodromal AD; (2) amyloid positivity had a spatially diffuse ALFF effect on frontal, medial parietal, and lateral temporal association cortices; (3) ERC ALFF, on the other hand, was associated with tau accumulation in the MTL; and, (4) all of these ALFF effects were observed in the absence of structural atrophy in preclinical AD. Together, these findings highlight ALFF as a measure that, at the group level, is more sensitive to AD pathology than macrostructural measurements in preclinical AD, supporting our hypothesis of earlier neurodegenerative changes in brain function.

While other work has suggested amyloid-related BOLD amplitude changes in preclinical AD (Millar et al., 2020), we also found local relationships with MTL tau in the absence of structural changes. We are aware of only one other study that evaluated the relationship between the amplitude of spontaneous BOLD signal fluctuations and tau in preclinical AD (Millar et al., 2020). That study investigated the relationship between resting-state BOLD variability (a measure similar to ALFF) and CSF pTau-181. The authors did not observe any statistically significant associations in preclinical AD. We suspect that tau PET and accurate MTL segmentations provided the anatomic specificity that is necessary to detect tau-driven ALFF effects in preclinical AD. Indeed, the most robust association between ALFF and tau was observed in the entorhinal cortex, a region that is particularly vulnerable to tau accumulation in the earliest stages of AD (Braak et al., 2006; Braak & Braak, 1991). Furthermore, since the effect of MTL tau on ERC ALFF was present even in the amyloid-negative control group, our results suggest that MTL ALFF might be sensitive to the tau accumulation of primary age-related tauopathy (PART), although some have argued that the signal in amyloid negative individuals may reflect non-specific neurodegeneration (Crary et al., 2014; Hickman et al., 2020; Landau & Mormino, 2023; Quintas-Neves et al., 2019).

Similarly, there has been little work on extra-MTL BOLD variability or ALFF differences in preclinical AD. Millar et al. (2020) reported trend-level associations between CSF Aβ42 and BOLD variability across a wide range of brain networks in CU older adults. In follow-up multivariate analyses, the authors showed that BOLD variability from brain areas belonging to the so-called default mode network (DMN), which includes medial frontal, precuneus, and posterior cingulate cortices, attained the best performance for predicting CSF Aβ42. Several studies also compared ALFF in cognitively normal older individuals with subjective cognitive decline to those without (Gao et al., 2023; Sun et al., 2016; Yang et al., 2018; Zhang et al., 2021); however, since no biomarker-based tests were used to ensure that those participants had AD pathology, it was not clear to what extent the reported ALFF differences were driven by AD as opposed to other neurodegenerative or non-neurodegenerative processes. In the present study, we showed that preclinical AD is associated with reduced ALFF in large segments of frontal, parietal, and temporal association cortices, as well as in the thalamus and striatum. These ALFF effects could not be explained by AD-associated cortical atrophy, and their spatial topography is consistent with well-established regions of amyloid accumulation in early AD (Therriault et al., 2022).

Previous ALFF studies in patients with aMCI led to inconsistent findings, with some studies reporting both increased and decreased ALFF in aMCI, sometimes in overlapping brain regions (Avants et al., 2019; Cha et al., 2015; Han et al., 2011; Liu et al., 2014; Tang et al., 2024; Xi et al., 2013). In general, those earlier studies showed MCI-associated ALFF reduction in the medial parietal cortex and insula and MCI-associated ALFF increase in the MTL and occipital regions (Pan et al., 2017). Our current results are in agreement with a longitudinal study by Avants et al. (2019), which demonstrated divergent spontaneous brain activity (adjusted measure of ALFF) patterns in Aβ-positive vs. Aβ-negative MCI over the 2-year follow-up window. The strongest statistical effects of Aβ were localized within the posterior segments of the default mode network, including the inferolateral parietal and precuneus cortices. After comparing patients with prodromal AD to age-matched controls without AD pathology, we did not observe a single brain region with increased ALFF in prodromal AD, regardless of the adjustment for GM atrophy. The incongruence of our current results with those from earlier studies might have been caused by differences in preprocessing pipelines, physiological noise removal techniques, and participant selection criteria.

Our mediation analyses revealed that the effects of amyloid and tau on ALFF are likely independent and complimentary. In the parietal cortex, amyloid positivity had a direct negative effect on ALFF, indicating that ALFF is sensitive to amyloid accumulation in the absence of tau. In the MTL, on the other hand, the effect of amyloid on ALFF was driven almost entirely by MTL tau, with ALFF progressively declining as the disease advanced. In our previous resting-state functional connectivity study (Hrybouski et al., 2023), we showed that functional connectivity in the anterior MTL is characterized by an ‘inverted U-shaped’ pattern with an initial rise during the preclinical stage followed by a decline in symptomatic AD. Though seemingly contradictory, these two sets of findings represent largely orthogonal measures: signal coherence between different brain areas and region-specific BOLD signal amplitude over time. It is thus plausible that the preclinical phase of AD is characterized by both widespread ALFF reduction in the neocortex and local hyperconnectivity centered around the anterior MTL. During the symptomatic stage of the disease, both ALFF and functional connectivity are negatively affected by disease progression (Hedden et al., 2009; Hrybouski et al., 2023).

The biological mechanisms underlying tau and amyloid effects on ALFF have yet to be elucidated. ALFF-like measures have weak-to-moderate correlations with cerebral blood flow (CBF) and ^18^F-fluoro-2-deoxyglucose (FDG) PET (Wang et al., 2021). Symptomatic AD is frequently associated with parietal and temporal hypometabolism on ^18^F-FDG PET (Chételat et al., 2020; Hanseeuw et al., 2019; Hoffman et al., 2000; Minoshima et al., 2022), and parietal hypometabolism has also been reported in Aβ-positive asymptomatic individuals with preclinical AD (Kljajevic et al., 2014). Because BOLD fMRI fluctuations over time are tightly coupled to metabolic demands of synaptic activity (Logothetis et al., 2001), reduced ALFF in AD might be driven by the loss of synapses or neurons in early disease that are not detectable with traditional macrostructural imaging. Because BOLD fMRI is neurovascular in nature, it is also plausible that reduced ALFF in AD is driven by pathologic cerebrovascular alterations, of which cerebral amyloid angiopathy (CAA) is particularly prominent (Merlini et al., 2016; Sweeney et al., 2018). In murine experiments, both amyloid and tau can induce hypoperfusion, potentially, in part, via abnormal remodeling of brain vasculature (Bennett et al., 2018; Klohs et al., 2014; Lourenço et al., 2017; Park et al., 2020). Conversely, hypoperfusion may also lead to an increase in amyloid and tau (Chan et al., 2018; Liang et al., 2015; Raz et al., 2019; Zhang et al., 2010). Negative effects of AD pathology on CBF have also been observed in human neuroimaging studies (Albrecht et al., 2020; Dolui et al., 2020; McDade et al., 2014; Wolk & Detre, 2012; Yan et al., 2018). Building on this earlier work, we showed that ALFF was a marginal mediator of the effect of amyloid on MTL tau, providing evidence that neurovascular abnormalities in early AD likely facilitate tau accumulation in the MTL.

### Limitations and future work

Resting-state functional MRI measures, including ALFF, are susceptible to motion-related artifacts (Satterthwaite et al., 2012). Previous studies showed that older adults tend to move more during MRI scanning than younger individuals (Hrybouski et al., 2021; Madan, 2018). Although we used a customized analysis pipeline to minimize the effect of such confounds on our ALFF measures, it is important to acknowledge that postprocessing-based denoising is not completely effective (Ciric et al., 2018; Satterthwaite et al., 2019). However, because head movement summary metrics were similar in all groups (see Suppl. Table 5), we do not think that residual head motion artifacts had a significant impact on our main findings.

The ALFF metric is prone to contamination by physiological noise (Zuo et al., 2010). To reduce the impact of physiological signals on our ALFF measurements, we used spatial ICA and aCompCorr techniques to identify and regress out confounding physiological signals from our fMRI data (Beckmann & Smith, 2004; Behzadi et al., 2007; Griffanti et al., 2017). Our fMRI acquisitions also featured sub-second temporal resolution, which also improves the effectiveness of temporal filters at removing physiological noise.

Although we demonstrated significant effects of AD pathology on MTL and cortical ALFF, it is not clear whether these effects are neural or vascular in origin. To definitively determine the biological origin of ALFF decline in preclinical and prodromal AD, quantitative fMRI or a combination of multimodal imaging techniques – including CBF, fMRI-based ALFF, and ^18^F-FDG PET – is necessary, and this would be a promising avenue for future research.

As with most BOLD fMRI studies, the anterior MTL subregions in the current study were more vulnerable to susceptibility artifacts than their posterior counterparts. However, it is unlikely that susceptibility artifacts biased our main findings because temporal signal-to-noise ratio profiles of ASHS-T1 ROIs were similar across groups (Suppl. Table 6). We anticipated that the relationship between tau and ALFF in BA35 would be comparable to that in ERC, given that both regions are vulnerable to tau accumulation in early Alzheimer’s disease (Braak & Braak, 1991). However, the tau-ALFF associations in BA35 did not reach statistical significance. It is currently unclear whether these differences are caused by physiological processes or by the varying susceptibility of the two regions to signal dropout in gradient echo EPI. Future research employing multi-echo acquisition protocols, which are less affected by such artifacts, will be able to directly compare the effects of AD pathology on ALFF in BA35 and ERC. Lastly, it is important to note that our study was cross-sectional. The extent to which our current results are representative of longitudinal trajectories along the AD continuum is not known.

## CONCLUSION

Our results indicate that Alzheimer’s pathology adversely affects both cortical and subcortical BOLD ALFF, even before detectable macrostructural change occurs in preclinical AD. Similarly, BOLD ALFF changes in prodromal AD are not driven by macrostructural atrophy. In the earliest stages of the disease, amyloid positivity is linked to a widespread reduction in ALFF, while tau accumulation specifically reduces ALFF in the medial temporal regions, particularly the entorhinal cortex. These findings suggest that changes in brain function occur earlier than detectable macrostructural changes and highlight the sensitivity of BOLD ALFF to early pathological changes in Alzheimer’s disease. Consequently, ALFF may be a promising metric in the context of amyloid-clearing interventions in preclinical Alzheimer’s disease.

## Data Availability

Anonymized pre-processed data and in-house analysis scripts will be made available upon request for the sole purpose of replicating procedures and results presented in this article.

## REFERENCES

Abramov, E., Dolev, I., Fogel, H., Ciccotosto, G. D., Ruff, E., & Slutsky, I. (2009). Amyloid-beta as a positive endogenous regulator of release probability at hippocampal synapses. Nat Neurosci, 12(12), 1567–1576. 10.1038/nn.2433

Adams, J. N., Maass, A., Berron, D., Harrison, T. M., Baker, S. L., Thomas, W. P., Stanfill, M., & Jagust, W. J. (2021). Reduced Repetition Suppression in Aging is Driven by Tau-Related Hyperactivity in Medial Temporal Lobe. J Neurosci, 41(17), 3917–3931. 10.1523/JNEUROSCI.2504-20.2021

Albrecht, D., Isenberg, A. L., Stradford, J., Monreal, T., Sagare, A., Pachicano, M., Sweeney, M., Toga, A., Zlokovic, B., Chui, H., Joe, E., Schneider, L., Conti, P., Jann, K., & Pa, J. (2020). Associations between Vascular Function and Tau PET Are Associated with Global Cognition and Amyloid. J Neurosci, 40(44), 8573–8586. 10.1523/JNEUROSCI.1230-20.2020

Alexopoulos, P., Sorg, C., Forschler, A., Grimmer, T., Skokou, M., Wohlschlager, A., Perneczky, R., Zimmer, C., Kurz, A., & Preibisch, C. (2012). Perfusion abnormalities in mild cognitive impairment and mild dementia in Alzheimer’s disease measured by pulsed arterial spin labeling MRI. Eur Arch Psychiatry Clin Neurosci, 262(1), 69–77. 10.1007/s00406-011-0226-2

Alsop, D. C., Detre, J. A., & Grossman, M. (2000). Assessment of cerebral blood flow in Alzheimer’s disease by spin-labeled magnetic resonance imaging. Ann Neurol, 47(1), 93–100. https://www.ncbi.nlm.nih.gov/pubmed/10632106

Angulo, S. L., Orman, R., Neymotin, S. A., Liu, L., Buitrago, L., Cepeda-Prado, E., Stefanov, D., Lytton, W. W., Stewart, M., Small, S. A., Duff, K. E., & Moreno, H. (2017). Tau and amyloid-related pathologies in the entorhinal cortex have divergent effects in the hippocampal circuit. Neurobiol Dis, 108, 261–276. 10.1016/j.nbd.2017.08.015

Asman, A. J., & Landman, B. A. (2013). Non-local statistical label fusion for multi-atlas segmentation. Medical Image Analysis, 17(2), 194–208. 10.1016/j.media.2012.10.002

Avants, B., & Gee, J. C. (2004). Geodesic estimation for large deformation anatomical shape averaging and interpolation. Neuroimage, 23 *Suppl 1*, S139–150. 10.1016/j.neuroimage.2004.07.010

Avants, B. B., Epstein, C. L., Grossman, M., & Gee, J. C. (2008). Symmetric diffeomorphic image registration with cross-correlation: evaluating automated labeling of elderly and neurodegenerative brain. Med Image Anal, 12(1), 26–41. 10.1016/j.media.2007.06.004

Avants, B. B., Hutchison, R. M., Mikulskis, A., Salinas-Valenzuela, C., Hargreaves, R., Beaver, J., Chiao, P., & Alzheimer’s Disease Neuroimaging, I. (2019). Amyloid beta-positive subjects exhibit longitudinal network-specific reductions in spontaneous brain activity. Neurobiol Aging, 74, 191–201. 10.1016/j.neurobiolaging.2018.10.002

Baller, E. B., Valcarcel, A. M., Adebimpe, A., Alexander-Bloch, A., Cui, Z., Gur, R. C., Gur, R. E., Larsen, B. L., Linn, K. A., O’Donnell, C. M., Pines, A. R., Raznahan, A., Roalf, D. R., Sydnor, V. J., Tapera, T. M., Tisdall, M. D., Vandekar, S., Xia, C. H., Detre, J. A., … Satterthwaite, T. D. (2022). Developmental coupling of cerebral blood flow and fMRI fluctuations in youth. Cell Rep, 38(13), 110576. 10.1016/j.celrep.2022.110576

Beckmann, C. F., & Smith, S. M. (2004). Probabilistic independent component analysis for functional magnetic resonance imaging. IEEE Trans Med Imaging, 23(2), 137–152. 10.1109/TMI.2003.822821

Behzadi, Y., Restom, K., Liau, J., & Liu, T. T. (2007). A component based noise correction method (CompCor) for BOLD and perfusion based fMRI. Neuroimage, 37(1), 90–101. 10.1016/j.neuroimage.2007.04.042

Bennett, R. E., Robbins, A. B., Hu, M., Cao, X., Betensky, R. A., Clark, T., Das, S., & Hyman, B. T. (2018). Tau induces blood vessel abnormalities and angiogenesis-related gene expression in P301L transgenic mice and human Alzheimer’s disease. Proc Natl Acad Sci U S A, 115(6), E1289–E1298. 10.1073/pnas.1710329115

Berron, D., van Westen, D., Ossenkoppele, R., Strandberg, O., & Hansson, O. (2020). Medial temporal lobe connectivity and its associations with cognition in early Alzheimer’s disease. Brain, 143(4), 1233–1248. 10.1093/brain/awaa068

Berron, D., Vogel, J. W., Insel, P. S., Pereira, J. B., Xie, L., Wisse, L. E. M., Yushkevich, P. A., Palmqvist, S., Mattsson-Carlgren, N., Stomrud, E., Smith, R., Strandberg, O., & Hansson, O. (2021). Early stages of tau pathology and its associations with functional connectivity, atrophy and memory. Brain, 144(9), 2771–2783. 10.1093/brain/awab114

Braak, H., Alafuzoff, I., Arzberger, T., Kretzschmar, H., & Del Tredici, K. (2006). Staging of Alzheimer disease-associated neurofibrillary pathology using paraffin sections and immunocytochemistry. Acta Neuropathol, 112(4), 389–404. 10.1007/s00401-006-0127-z

Braak, H., & Braak, E. (1991). Neuropathological stageing of Alzheimer-related changes. Acta Neuropathol, 82(4), 239–259. 10.1007/BF00308809

Braun, U., Plichta, M. M., Esslinger, C., Sauer, C., Haddad, L., Grimm, O., Mier, D., Mohnke, S., Heinz, A., Erk, S., Walter, H., Seiferth, N., Kirsch, P., & Meyer-Lindenberg, A. (2012). Test-retest reliability of resting-state connectivity network characteristics using fMRI and graph theoretical measures. Neuroimage, 59(2), 1404–1412. 10.1016/j.neuroimage.2011.08.044

Buckner, R. L., Snyder, A. Z., Shannon, B. J., LaRossa, G., Sachs, R., Fotenos, A. F., Sheline, Y. I., Klunk, W. E., Mathis, C. A., Morris, J. C., & Mintun, M. A. (2005). Molecular, structural, and functional characterization of Alzheimer’s disease: evidence for a relationship between default activity, amyloid, and memory. J Neurosci, 25(34), 7709–7717. 10.1523/JNEUROSCI.2177-05.2005

Busche, M. A., Chen, X., Henning, H. A., Reichwald, J., Staufenbiel, M., Sakmann, B., & Konnerth, A. (2012). Critical role of soluble amyloid-beta for early hippocampal hyperactivity in a mouse model of Alzheimer’s disease. Proc Natl Acad Sci U S A, 109(22), 8740–8745. 10.1073/pnas.1206171109

Cahart, M. S., O’Daly, O., Giampietro, V., Timmers, M., Streffer, J., Einstein, S., Zelaya, F., Dell’Acqua, F., & Williams, S. C. R. (2023). Comparing the test-retest reliability of resting-state functional magnetic resonance imaging metrics across single band and multiband acquisitions in the context of healthy aging. Hum Brain Mapp, 44(5), 1901–1912. 10.1002/hbm.26180

Cha, J., Hwang, J. M., Jo, H. J., Seo, S. W., Na, D. L., & Lee, J. M. (2015). Assessment of Functional Characteristics of Amnestic Mild Cognitive Impairment and Alzheimer’s Disease Using Various Methods of Resting-State FMRI Analysis. Biomed Res Int, 2015, 907464. 10.1155/2015/907464

Chan, S. L., Bishop, N., Li, Z., & Cipolla, M. J. (2018). Inhibition of PAI (Plasminogen Activator Inhibitor)-1 Improves Brain Collateral Perfusion and Injury After Acute Ischemic Stroke in Aged Hypertensive Rats. Stroke, 49(8), 1969–1976. 10.1161/STROKEAHA.118.022056

Chételat, G., Arbizu, J., Barthel, H., Garibotto, V., Law, I., Morbelli, S., van de Giessen, E., Agosta, F., Barkhof, F., Brooks, D. J., Carrillo, M. C., Dubois, B., Fjell, A. M., Frisoni, G. B., Hansson, O., Herholz, K., Hutton, B. F., Jack, C. R., Jr., Lammertsma, A. A., … Drzezga, A. (2020). Amyloid-PET and (18)F-FDG-PET in the diagnostic investigation of Alzheimer’s disease and other dementias. Lancet Neurol, 19(11), 951–962. 10.1016/S1474-4422(20)30314-8

Chételat, G., Desgranges, B., de la Sayette, V., Viader, F., Eustache, F., & Baron, J. C. (2003). Mild cognitive impairment: Can FDG-PET predict who is to rapidly convert to Alzheimer’s disease? Neurology, 60(8), 1374–1377. 10.1212/01.wnl.0000055847.17752.e6

Cho, H., Choi, J. Y., Hwang, M. S., Kim, Y. J., Lee, H. M., Lee, H. S., Lee, J. H., Ryu, Y. H., Lee, M. S., & Lyoo, C. H. (2016). In vivo cortical spreading pattern of tau and amyloid in the Alzheimer disease spectrum. Ann Neurol, 80(2), 247–258. 10.1002/ana.24711

Ciric, R., Rosen, A. F. G., Erus, G., Cieslak, M., Adebimpe, A., Cook, P. A., Bassett, D. S., Davatzikos, C., Wolf, D. H., & Satterthwaite, T. D. (2018). Mitigating head motion artifact in functional connectivity MRI. Nat Protoc, 13(12), 2801–2826. 10.1038/s41596-018-0065-y

Cox, R. W., & Hyde, J. S. (1997). Software tools for analysis and visualization of fMRI data. NMR Biomed, 10(4-5), 171–178. 10.1002/(sici)1099-1492(199706/08)10:4/5<171::aid-nbm453>3.0.co;2-l

Crary, J. F., Trojanowski, J. Q., Schneider, J. A., Abisambra, J. F., Abner, E. L., Alafuzoff, I., Arnold, S. E., Attems, J., Beach, T. G., Bigio, E. H., Cairns, N. J., Dickson, D. W., Gearing, M., Grinberg, L. T., Hof, P. R., Hyman, B. T., Jellinger, K., Jicha, G. A., Kovacs, G. G., … Nelson, P. T. (2014). Primary age-related tauopathy (PART): a common pathology associated with human aging. Acta Neuropathol, 128(6), 755–766. 10.1007/s00401-014-1349-0

Das, S. R., Avants, B. B., Grossman, M., & Gee, J. C. (2009). Registration based cortical thickness measurement. Neuroimage, 45(3), 867–879. 10.1016/j.neuroimage.2008.12.016

Das, S. R., Pluta, J., Mancuso, L., Kliot, D., Orozco, S., Dickerson, B. C., Yushkevich, P. A., & Wolk, D. A. (2013). Increased functional connectivity within medial temporal lobe in mild cognitive impairment. Hippocampus, 23(1), 1–6. 10.1002/hipo.22051

Dolui, S., Li, Z., Nasrallah, I. M., Detre, J. A., & Wolk, D. A. (2020). Arterial spin labeling versus (18)F-FDG-PET to identify mild cognitive impairment. Neuroimage Clin, 25, 102146. 10.1016/j.nicl.2019.102146

Elman, J. A., Madison, C. M., Baker, S. L., Vogel, J. W., Marks, S. M., Crowley, S., O’Neil, J. P., & Jagust, W. J. (2016). Effects of Beta-Amyloid on Resting State Functional Connectivity Within and Between Networks Reflect Known Patterns of Regional Vulnerability. Cereb Cortex, 26(2), 695–707. 10.1093/cercor/bhu259

Esteban, O., Markiewicz, C. J., Blair, R. W., Moodie, C. A., Isik, A. I., Erramuzpe, A., Kent, J. D., Goncalves, M., DuPre, E., Snyder, M., Oya, H., Ghosh, S. S., Wright, J., Durnez, J., Poldrack, R. A., & Gorgolewski, K. J. (2019). fMRIPrep: a robust preprocessing pipeline for functional MRI. Nat Methods, 16(1), 111–116. 10.1038/s41592-018-0235-4

Fair, D. A., Miranda-Dominguez, O., Snyder, A. Z., Perrone, A., Earl, E. A., Van, A. N., Koller, J. M., Feczko, E., Tisdall, M. D., van der Kouwe, A., Klein, R. L., Mirro, A. E., Hampton, J. M., Adeyemo, B., Laumann, T. O., Gratton, C., Greene, D. J., Schlaggar, B. L., Hagler, D. J., Jr., … Dosenbach, N. U. F. (2020). Correction of respiratory artifacts in MRI head motion estimates. Neuroimage, 208, 116400. 10.1016/j.neuroimage.2019.116400

Freedman, D., & Lane, D. (1983). A Nonstochastic Interpretation of Reported Significance Levels. Journal of Business & Economic Statistics, 1(4), 292–298. 10.2307/1391660

Friedland, R. P., Jagust, W. J., Huesman, R. H., Koss, E., Knittel, B., Mathis, C. A., Ober, B. A., Mazoyer, B. M., & Budinger, T. F. (1989). Regional cerebral glucose transport and utilization in Alzheimer’s disease. Neurology, 39(11), 1427–1434. 10.1212/wnl.39.11.1427

Friston, K. J., Williams, S., Howard, R., Frackowiak, R. S., & Turner, R. (1996). Movement-related effects in fMRI time-series. Magn Reson Med, 35(3), 346–355. 10.1002/mrm.1910350312

Gao, Y., Tian, S., Tang, Y., Yang, X., Dou, W., Wang, T., Shen, Y., Tang, Y., Zhang, L., Ding, H., Zhu, Q., Li, J., Qi, M., & Zhu, Y. (2023). Investigating the spontaneous brain activities of patients with subjective cognitive decline and mild cognitive impairment: an amplitude of low-frequency fluctuation functional magnetic resonance imaging study. Quant Imaging Med Surg, 13(12), 8557–8570. 10.21037/qims-23-808

Gorgolewski, K., Burns, C. D., Madison, C., Clark, D., Halchenko, Y. O., Waskom, M. L., & Ghosh, S. S. (2011). Nipype: a flexible, lightweight and extensible neuroimaging data processing framework in python. Front Neuroinform, 5, 13. 10.3389/fninf.2011.00013

Gratton, C., Dworetsky, A., Coalson, R. S., Adeyemo, B., Laumann, T. O., Wig, G. S., Kong, T. S., Gratton, G., Fabiani, M., Barch, D. M., Tranel, D., Miranda-Dominguez, O., Fair, D. A., Dosenbach, N. U. F., Snyder, A. Z., Perlmutter, J. S., Petersen, S. E., & Campbell, M. C. (2020). Removal of high frequency contamination from motion estimates in single-band fMRI saves data without biasing functional connectivity. Neuroimage, 217, 116866. 10.1016/j.neuroimage.2020.116866

Greve, D. N., & Fischl, B. (2009). Accurate and robust brain image alignment using boundary-based registration. Neuroimage, 48(1), 63–72. 10.1016/j.neuroimage.2009.06.060

Griffanti, L., Douaud, G., Bijsterbosch, J., Evangelisti, S., Alfaro-Almagro, F., Glasser, M. F., Duff, E. P., Fitzgibbon, S., Westphal, R., Carone, D., Beckmann, C. F., & Smith, S. M. (2017). Hand classification of fMRI ICA noise components. Neuroimage, 154, 188–205. 10.1016/j.neuroimage.2016.12.036

Han, Y., Wang, J., Zhao, Z., Min, B., Lu, J., Li, K., He, Y., & Jia, J. (2011). Frequency-dependent changes in the amplitude of low-frequency fluctuations in amnestic mild cognitive impairment: a resting-state fMRI study. Neuroimage, 55(1), 287–295. 10.1016/j.neuroimage.2010.11.059

Hanseeuw, B. J., Betensky, R. A., Jacobs, H. I. L., Schultz, A. P., Sepulcre, J., Becker, J. A., Cosio, D. M. O., Farrell, M., Quiroz, Y. T., Mormino, E. C., Buckley, R. F., Papp, K. V., Amariglio, R. A., Dewachter, I., Ivanoiu, A., Huijbers, W., Hedden, T., Marshall, G. A., Chhatwal, J. P., … Johnson, K. (2019). Association of Amyloid and Tau With Cognition in Preclinical Alzheimer Disease: A Longitudinal Study. JAMA Neurol, 76(8), 915–924. 10.1001/jamaneurol.2019.1424

Hedden, T., Van Dijk, K. R., Becker, J. A., Mehta, A., Sperling, R. A., Johnson, K. A., & Buckner, R. L. (2009). Disruption of functional connectivity in clinically normal older adults harboring amyloid burden. J Neurosci, 29(40), 12686–12694. 10.1523/JNEUROSCI.3189-09.2009

Hickman, R. A., Flowers, X. E., & Wisniewski, T. (2020). Primary Age-Related Tauopathy (PART): Addressing the Spectrum of Neuronal Tauopathic Changes in the Aging Brain. Curr Neurol Neurosci Rep, 20(9), 39. 10.1007/s11910-020-01063-1

Hoffman, J. M., Welsh-Bohmer, K. A., Hanson, M., Crain, B., Hulette, C., Earl, N., & Coleman, R. E. (2000). FDG PET imaging in patients with pathologically verified dementia. J Nucl Med, 41(11), 1920–1928. https://www.ncbi.nlm.nih.gov/pubmed/11079505

Hrybouski, S., Cribben, I., McGonigle, J., Olsen, F., Carter, R., Seres, P., Madan, C. R., & Malykhin, N. V. (2021). Investigating the effects of healthy cognitive aging on brain functional connectivity using 4.7 T resting-state functional magnetic resonance imaging. Brain Struct Funct, 226(4), 1067–1098. 10.1007/s00429-021-02226-7

Hrybouski, S., Das, S. R., Xie, L., Wisse, L. E. M., Kelley, M., Lane, J., Sherin, M., DiCalogero, M., Nasrallah, I., Detre, J., Yushkevich, P. A., & Wolk, D. A. (2023). Aging and Alzheimer’s disease have dissociable effects on local and regional medial temporal lobe connectivity. Brain Commun, 5(5), fcad245. 10.1093/braincomms/fcad245

Ishii, K., Sasaki, M., Yamaji, S., Sakamoto, S., Kitagaki, H., & Mori, E. (1997). Demonstration of decreased posterior cingulate perfusion in mild Alzheimer’s disease by means of H215O positron emission tomography. Eur J Nucl Med, 24(6), 670–673. 10.1007/BF00841407

Jack, C. R., Jr., Andrews, J. S., Beach, T. G., Buracchio, T., Dunn, B., Graf, A., Hansson, O., Ho, C., Jagust, W., McDade, E., Molinuevo, J. L., Okonkwo, O. C., Pani, L., Rafii, M. S., Scheltens, P., Siemers, E., Snyder, H. M., Sperling, R., Teunissen, C. E., & Carrillo, M. C. (2024). Revised criteria for diagnosis and staging of Alzheimer’s disease: Alzheimer’s Association Workgroup. Alzheimers Dement. 10.1002/alz.13859

Jenkinson, M., Bannister, P., Brady, M., & Smith, S. (2002). Improved optimization for the robust and accurate linear registration and motion correction of brain images. Neuroimage, 17(2), 825–841. 10.1016/s1053-8119(02)91132-8

Jezzard, P., & Balaban, R. S. (1995). Correction for geometric distortion in echo planar images from B0 field variations. Magn Reson Med, 34(1), 65–73. 10.1002/mrm.1910340111

Kenny, D. A., Korchmaros, J. D., & Bolger, N. (2003). Lower level mediation in multilevel models. Psychol Methods, 8(2), 115–128. 10.1037/1082-989x.8.2.115

Klein, A., & Tourville, J. (2012). 101 labeled brain images and a consistent human cortical labeling protocol. Front Neurosci, 6, 171. 10.3389/fnins.2012.00171

Kljajevic, V., Grothe, M. J., Ewers, M., Teipel, S., & Alzheimer’s Disease Neuroimaging, I. (2014). Distinct pattern of hypometabolism and atrophy in preclinical and predementia Alzheimer’s disease. Neurobiol Aging, 35(9), 1973–1981. 10.1016/j.neurobiolaging.2014.04.006

Klohs, J., Rudin, M., Shimshek, D. R., & Beckmann, N. (2014). Imaging of cerebrovascular pathology in animal models of Alzheimer’s disease. Front Aging Neurosci, 6, 32. 10.3389/fnagi.2014.00032

Landau, S. M., Breault, C., Joshi, A. D., Pontecorvo, M., Mathis, C. A., Jagust, W. J., Mintun, M. A., & Alzheimer’s Disease Neuroimaging, I. (2013). Amyloid-beta imaging with Pittsburgh compound B and florbetapir: comparing radiotracers and quantification methods. J Nucl Med, 54(1), 70–77. 10.2967/jnumed.112.109009

Landau, S. M., & Mormino, E. C. (2023). Tau Pathology Without Abeta-A Limited PART of Clinical Progression. JAMA Neurol, 80(10), 1025–1027. 10.1001/jamaneurol.2023.1081

Landman, B., & Warfield, S. (2012). MICCAI 2012 workshop on multi-atlas labeling. In MICCAI Grand Challenge and Workshop on Multi-Atlas Labeling. CreateSpace Independent Publishing Platform.

Lemieux, L., Salek-Haddadi, A., Lund, T. E., Laufs, H., & Carmichael, D. (2007). Modelling large motion events in fMRI studies of patients with epilepsy. Magn Reson Imaging, 25(6), 894–901. 10.1016/j.mri.2007.03.009

Liang, W., Zhang, W., Zhao, S., Li, Q., Liang, H., & Ceng, R. (2015). Altered expression of neurofilament 200 and amyloid-beta peptide (1-40) in a rat model of chronic cerebral hypoperfusion. Neurol Sci, 36(5), 707–712. 10.1007/s10072-014-2014-z

Liu, X., Wang, S., Zhang, X., Wang, Z., Tian, X., & He, Y. (2014). Abnormal amplitude of low-frequency fluctuations of intrinsic brain activity in Alzheimer’s disease. J Alzheimers Dis, 40(2), 387–397. 10.3233/JAD-131322

Logothetis, N. K., Pauls, J., Augath, M., Trinath, T., & Oeltermann, A. (2001). Neurophysiological investigation of the basis of the fMRI signal. Nature, 412(6843), 150–157. 10.1038/35084005

Lourenço, C. F., Ledo, A., Barbosa, R. M., & Laranjinha, J. (2017). Neurovascular uncoupling in the triple transgenic model of Alzheimer’s disease: Impaired cerebral blood flow response to neuronal-derived nitric oxide signaling. Exp Neurol, 291, 36–43. 10.1016/j.expneurol.2017.01.013

Maass, A., Berron, D., Harrison, T. M., Adams, J. N., La Joie, R., Baker, S., Mellinger, T., Bell, R. K., Swinnerton, K., Inglis, B., Rabinovici, G. D., Duzel, E., & Jagust, W. J. (2019). Alzheimer’s pathology targets distinct memory networks in the ageing brain. Brain, 142(8), 2492–2509. 10.1093/brain/awz154

Madan, C. R. (2018). Age differences in head motion and estimates of cortical morphology. PeerJ, 6, e5176. 10.7717/peerj.5176

Mattsson, N., Tosun, D., Insel, P. S., Simonson, A., Jack, C. R., Jr., Beckett, L. A., Donohue, M., Jagust, W., Schuff, N., Weiner, M. W., & Alzheimer’s Disease Neuroimaging, I. (2014). Association of brain amyloid-beta with cerebral perfusion and structure in Alzheimer’s disease and mild cognitive impairment. Brain, 137(Pt 5), 1550–1561. 10.1093/brain/awu043

McDade, E., Kim, A., James, J., Sheu, L. K., Kuan, D. C., Minhas, D., Gianaros, P. J., Ikonomovic, S., Lopez, O., Snitz, B., Price, J., Becker, J., Mathis, C., & Klunk, W. (2014). Cerebral perfusion alterations and cerebral amyloid in autosomal dominant Alzheimer disease. Neurology, 83(8), 710–717. 10.1212/WNL.0000000000000721

Merlini, M., Wanner, D., & Nitsch, R. M. (2016). Tau pathology-dependent remodelling of cerebral arteries precedes Alzheimer’s disease-related microvascular cerebral amyloid angiopathy. Acta Neuropathol, 131(5), 737–752. 10.1007/s00401-016-1560-2

Millar, P. R., Ances, B. M., Gordon, B. A., Benzinger, T. L. S., Fagan, A. M., Morris, J. C., & Balota, D. A. (2020). Evaluating resting-state BOLD variability in relation to biomarkers of preclinical Alzheimer’s disease. Neurobiol Aging, 96, 233–245. 10.1016/j.neurobiolaging.2020.08.007

Minoshima, S., Cross, D., Thientunyakit, T., Foster, N. L., & Drzezga, A. (2022). (18)F-FDG PET Imaging in Neurodegenerative Dementing Disorders: Insights into Subtype Classification, Emerging Disease Categories, and Mixed Dementia with Copathologies. J Nucl Med, 63(Suppl 1), 2S–12S. 10.2967/jnumed.121.263194

Mormino, E. C., Smiljic, A., Hayenga, A. O., Onami, S. H., Greicius, M. D., Rabinovici, G. D., Janabi, M., Baker, S. L., Yen, I. V., Madison, C. M., Miller, B. L., & Jagust, W. J. (2011). Relationships between beta-amyloid and functional connectivity in different components of the default mode network in aging. Cereb Cortex, 21(10), 2399–2407. 10.1093/cercor/bhr025

Mosconi, L., Tsui, W. H., Herholz, K., Pupi, A., Drzezga, A., Lucignani, G., Reiman, E. M., Holthoff, V., Kalbe, E., Sorbi, S., Diehl-Schmid, J., Perneczky, R., Clerici, F., Caselli, R., Beuthien-Baumann, B., Kurz, A., Minoshima, S., & de Leon, M. J. (2008). Multicenter standardized 18F-FDG PET diagnosis of mild cognitive impairment, Alzheimer’s disease, and other dementias. J Nucl Med, 49(3), 390–398. 10.2967/jnumed.107.045385

Noble, S., Scheinost, D., & Constable, R. T. (2019). A decade of test-retest reliability of functional connectivity: A systematic review and meta-analysis. Neuroimage, 203, 116157. 10.1016/j.neuroimage.2019.116157

Noble, S., Spann, M. N., Tokoglu, F., Shen, X., Constable, R. T., & Scheinost, D. (2017). Influences on the Test-Retest Reliability of Functional Connectivity MRI and its Relationship with Behavioral Utility. Cereb Cortex, 27(11), 5415–5429. 10.1093/cercor/bhx230

Nugent, A. C., Martinez, A., D’Alfonso, A., Zarate, C. A., & Theodore, W. H. (2015). The relationship between glucose metabolism, resting-state fMRI BOLD signal, and GABAA-binding potential: a preliminary study in healthy subjects and those with temporal lobe epilepsy. J Cereb Blood Flow Metab, 35(4), 583–591. 10.1038/jcbfm.2014.228

Palmqvist, S., Scholl, M., Strandberg, O., Mattsson, N., Stomrud, E., Zetterberg, H., Blennow, K., Landau, S., Jagust, W., & Hansson, O. (2017). Earliest accumulation of beta-amyloid occurs within the default-mode network and concurrently affects brain connectivity. Nat Commun, 8(1), 1214. 10.1038/s41467-017-01150-x

Palop, J. J., & Mucke, L. (2016). Network abnormalities and interneuron dysfunction in Alzheimer disease. Nat Rev Neurosci, 17(12), 777–792. 10.1038/nrn.2016.141

Pan, P., Zhu, L., Yu, T., Shi, H., Zhang, B., Qin, R., Zhu, X., Qian, L., Zhao, H., Zhou, H., & Xu, Y. (2017). Aberrant spontaneous low-frequency brain activity in amnestic mild cognitive impairment: A meta-analysis of resting-state fMRI studies. Ageing Res Rev, 35, 12–21. 10.1016/j.arr.2016.12.001

Park, L., Hochrainer, K., Hattori, Y., Ahn, S. J., Anfray, A., Wang, G., Uekawa, K., Seo, J., Palfini, V., Blanco, I., Acosta, D., Eliezer, D., Zhou, P., Anrather, J., & Iadecola, C. (2020). Tau induces PSD95-neuronal NOS uncoupling and neurovascular dysfunction independent of neurodegeneration. Nat Neurosci, 23(9), 1079–1089. 10.1038/s41593-020-0686-7

Pooler, A. M., Phillips, E. C., Lau, D. H., Noble, W., & Hanger, D. P. (2013). Physiological release of endogenous tau is stimulated by neuronal activity. EMBO Rep, 14(4), 389–394. 10.1038/embor.2013.15

Power, J. D., Mitra, A., Laumann, T. O., Snyder, A. Z., Schlaggar, B. L., & Petersen, S. E. (2014). Methods to detect, characterize, and remove motion artifact in resting state fMRI. Neuroimage, 84, 320–341. 10.1016/j.neuroimage.2013.08.048

Pruim, R. H. R., Mennes, M., van Rooij, D., Llera, A., Buitelaar, J. K., & Beckmann, C. F. (2015). ICA-AROMA: A robust ICA-based strategy for removing motion artifacts from fMRI data. Neuroimage, 112, 267–277. 10.1016/j.neuroimage.2015.02.064

Quintas-Neves, M., Teylan, M. A., Besser, L., Soares-Fernandes, J., Mock, C. N., Kukull, W. A., Crary, J. F., & Oliveira, T. G. (2019). Magnetic resonance imaging brain atrophy assessment in primary age-related tauopathy (PART). Acta Neuropathol Commun, 7(1), 204. 10.1186/s40478-019-0842-z

Raichle, M. E., MacLeod, A. M., Snyder, A. Z., Powers, W. J., Gusnard, D. A., & Shulman, G. L. (2001). A default mode of brain function. Proc Natl Acad Sci U S A, 98(2), 676–682. 10.1073/pnas.98.2.676

Raz, L., Bhaskar, K., Weaver, J., Marini, S., Zhang, Q., Thompson, J. F., Espinoza, C., Iqbal, S., Maphis, N. M., Weston, L., Sillerud, L. O., Caprihan, A., Pesko, J. C., Erhardt, E. B., & Rosenberg, G. A. (2019). Hypoxia promotes tau hyperphosphorylation with associated neuropathology in vascular dysfunction. Neurobiol Dis, 126, 124–136. 10.1016/j.nbd.2018.07.009

Satterthwaite, T. D., Ciric, R., Roalf, D. R., Davatzikos, C., Bassett, D. S., & Wolf, D. H. (2019). Motion artifact in studies of functional connectivity: Characteristics and mitigation strategies. Hum Brain Mapp, 40(7), 2033–2051. 10.1002/hbm.23665

Satterthwaite, T. D., Elliott, M. A., Gerraty, R. T., Ruparel, K., Loughead, J., Calkins, M. E., Eickhoff, S. B., Hakonarson, H., Gur, R. C., Gur, R. E., & Wolf, D. H. (2013). An improved framework for confound regression and filtering for control of motion artifact in the preprocessing of resting-state functional connectivity data. Neuroimage, 64, 240–256. 10.1016/j.neuroimage.2012.08.052

Satterthwaite, T. D., Wolf, D. H., Loughead, J., Ruparel, K., Elliott, M. A., Hakonarson, H., Gur, R. C., & Gur, R. E. (2012). Impact of in-scanner head motion on multiple measures of functional connectivity: relevance for studies of neurodevelopment in youth. Neuroimage, 60(1), 623–632. 10.1016/j.neuroimage.2011.12.063

Schoonhoven, D. N., Coomans, E. M., Millan, A. P., van Nifterick, A. M., Visser, D., Ossenkoppele, R., Tuncel, H., van der Flier, W. M., Golla, S. S. V., Scheltens, P., Hillebrand, A., van Berckel, B. N. M., Stam, C. J., & Gouw, A. A. (2023). Tau protein spreads through functionally connected neurons in Alzheimer’s disease: a combined MEG/PET study. Brain, 146(10), 4040–4054. 10.1093/brain/awad189

Sheline, Y. I., Raichle, M. E., Snyder, A. Z., Morris, J. C., Head, D., Wang, S., & Mintun, M. A. (2010). Amyloid plaques disrupt resting state default mode network connectivity in cognitively normal elderly. Biol Psychiatry, 67(6), 584–587. 10.1016/j.biopsych.2009.08.024

Shrout, P. E., & Bolger, N. (2002). Mediation in experimental and nonexperimental studies: new procedures and recommendations. Psychol Methods, 7(4), 422–445. https://www.ncbi.nlm.nih.gov/pubmed/12530702

Silverman, D. H., Small, G. W., Chang, C. Y., Lu, C. S., Kung De Aburto, M. A., Chen, W., Czernin, J., Rapoport, S. I., Pietrini, P., Alexander, G. E., Schapiro, M. B., Jagust, W. J., Hoffman, J. M., Welsh-Bohmer, K. A., Alavi, A., Clark, C. M., Salmon, E., de Leon, M. J., Mielke, R., … Phelps, M. E. (2001). Positron emission tomography in evaluation of dementia: Regional brain metabolism and long-term outcome. JAMA, 286(17), 2120–2127. 10.1001/jama.286.17.2120

Sims, J. R., Zimmer, J. A., Evans, C. D., Lu, M., Ardayfio, P., Sparks, J., Wessels, A. M., Shcherbinin, S., Wang, H., Monkul Nery, E. S., Collins, E. C., Solomon, P., Salloway, S., Apostolova, L. G., Hansson, O., Ritchie, C., Brooks, D. A., Mintun, M., Skovronsky, D. M., & Investigators, T.-A. (2023). Donanemab in Early Symptomatic Alzheimer Disease: The TRAILBLAZER-ALZ 2 Randomized Clinical Trial. JAMA, 330(6), 512–527. 10.1001/jama.2023.13239

Smith, S. M., Jenkinson, M., Woolrich, M. W., Beckmann, C. F., Behrens, T. E., Johansen-Berg, H., Bannister, P. R., De Luca, M., Drobnjak, I., Flitney, D. E., Niazy, R. K., Saunders, J., Vickers, J., Zhang, Y., De Stefano, N., Brady, J. M., & Matthews, P. M. (2004). Advances in functional and structural MR image analysis and implementation as FSL. Neuroimage, 23 *Suppl 1*, S208–219. 10.1016/j.neuroimage.2004.07.051

Smith, S. M., & Nichols, T. E. (2009). Threshold-free cluster enhancement: addressing problems of smoothing, threshold dependence and localisation in cluster inference. Neuroimage, 44(1), 83–98. 10.1016/j.neuroimage.2008.03.061

Sojkova, J., Beason-Held, L., Zhou, Y., An, Y., Kraut, M. A., Ye, W., Ferrucci, L., Mathis, C. A., Klunk, W. E., Wong, D. F., & Resnick, S. M. (2008). Longitudinal cerebral blood flow and amyloid deposition: an emerging pattern? J Nucl Med, 49(9), 1465–1471. 10.2967/jnumed.108.051946

Sperling, R. A., Donohue, M. C., Raman, R., Rafii, M. S., Johnson, K., Masters, C. L., van Dyck, C. H., Iwatsubo, T., Marshall, G. A., Yaari, R., Mancini, M., Holdridge, K. C., Case, M., Sims, J. R., Aisen, P. S., & Team, A. S. (2023). Trial of Solanezumab in Preclinical Alzheimer’s Disease. N Engl J Med, 389(12), 1096–1107. 10.1056/NEJMoa2305032

Sperling, R. A., Laviolette, P. S., O’Keefe, K., O’Brien, J., Rentz, D. M., Pihlajamaki, M., Marshall, G., Hyman, B. T., Selkoe, D. J., Hedden, T., Buckner, R. L., Becker, J. A., & Johnson, K. A. (2009). Amyloid deposition is associated with impaired default network function in older persons without dementia. Neuron, 63(2), 178–188. 10.1016/j.neuron.2009.07.003

Stargardt, A., Swaab, D. F., & Bossers, K. (2015). Storm before the quiet: neuronal hyperactivity and Abeta in the presymptomatic stages of Alzheimer’s disease. Neurobiol Aging, 36(1), 1–11. 10.1016/j.neurobiolaging.2014.08.014

Sun, Y., Dai, Z., Li, Y., Sheng, C., Li, H., Wang, X., Chen, X., He, Y., & Han, Y. (2016). Subjective Cognitive Decline: Mapping Functional and Structural Brain Changes-A Combined Resting-State Functional and Structural MR Imaging Study. Radiology, 281(1), 185–192. 10.1148/radiol.2016151771

Sweeney, M. D., Kisler, K., Montagne, A., Toga, A. W., & Zlokovic, B. V. (2018). The role of brain vasculature in neurodegenerative disorders. Nat Neurosci, 21(10), 1318–1331. 10.1038/s41593-018-0234-x

Tang, X., Guo, Z., Chen, G., Sun, S., Xiao, S., Chen, P., Tang, G., Huang, L., & Wang, Y. (2024). A Multimodal Meta-Analytical Evidence of Functional and Structural Brain Abnormalities Across Alzheimer’s Disease Spectrum. Ageing Res Rev, 95, 102240. 10.1016/j.arr.2024.102240

Thal, D. R., Rub, U., Orantes, M., & Braak, H. (2002). Phases of A beta-deposition in the human brain and its relevance for the development of AD. Neurology, 58(12), 1791–1800. 10.1212/wnl.58.12.1791

Therriault, J., Pascoal, T. A., Lussier, F. Z., Tissot, C., Chamoun, M., Bezgin, G., Servaes, S., Benedet, A. L., Ashton, N. J., Karikari, T. K., Lantero-Rodriguez, J., Kunach, P., Wang, Y.-T., Fernandez-Arias, J., Massarweh, G., Vitali, P., Soucy, J.-P., Saha-Chaudhuri, P., Blennow, K., … Rosa-Neto, P. (2022). Biomarker modeling of Alzheimer’s disease using PET-based Braak staging. Nature Aging, 2(6), 526–535. 10.1038/s43587-022-00204-0

van Dyck, C. H., Swanson, C. J., Aisen, P., Bateman, R. J., Chen, C., Gee, M., Kanekiyo, M., Li, D., Reyderman, L., Cohen, S., Froelich, L., Katayama, S., Sabbagh, M., Vellas, B., Watson, D., Dhadda, S., Irizarry, M., Kramer, L. D., & Iwatsubo, T. (2023). Lecanemab in Early Alzheimer’s Disease. N Engl J Med, 388(1), 9–21. 10.1056/NEJMoa2212948

Villeneuve, S., Rabinovici, G. D., Cohn-Sheehy, B. I., Madison, C., Ayakta, N., Ghosh, P. M., La Joie, R., Arthur-Bentil, S. K., Vogel, J. W., Marks, S. M., Lehmann, M., Rosen, H. J., Reed, B., Olichney, J., Boxer, A. L., Miller, B. L., Borys, E., Jin, L. W., Huang, E. J., … Jagust, W. (2015). Existing Pittsburgh Compound-B positron emission tomography thresholds are too high: statistical and pathological evaluation. Brain, 138(Pt 7), 2020–2033. 10.1093/brain/awv112

Wager, T. D., Davidson, M. L., Hughes, B. L., Lindquist, M. A., & Ochsner, K. N. (2008). Prefrontal-subcortical pathways mediating successful emotion regulation. Neuron, 59(6), 1037–1050. 10.1016/j.neuron.2008.09.006

Wang, H., Suh, J. W., Das, S. R., Pluta, J. B., Craige, C., & Yushkevich, P. A. (2013). Multi-Atlas Segmentation with Joint Label Fusion. IEEE Trans Pattern Anal Mach Intell, 35(3), 611–623. 10.1109/TPAMI.2012.143

Wang, H., & Yushkevich, P. A. (2013). Multi-atlas segmentation with joint label fusion and corrective learning-an open source implementation. Front Neuroinform, 7, 27. 10.3389/fninf.2013.00027

Wang, J., Sun, H., Cui, B., Yang, H., Shan, Y., Dong, C., Zang, Y., & Lu, J. (2021). The Relationship Among Glucose Metabolism, Cerebral Blood Flow, and Functional Activity: a Hybrid PET/fMRI Study. Mol Neurobiol, 58(6), 2862–2873. 10.1007/s12035-021-02305-0

Wang, X., Jiao, Y., Tang, T., Wang, H., & Lu, Z. (2013). Investigating univariate temporal patterns for intrinsic connectivity networks based on complexity and low-frequency oscillation: a test-retest reliability study. Neuroscience, 254, 404–426. 10.1016/j.neuroscience.2013.09.009

Weintraub, S., Besser, L., Dodge, H. H., Teylan, M., Ferris, S., Goldstein, F. C., Giordani, B., Kramer, J., Loewenstein, D., Marson, D., Mungas, D., Salmon, D., Welsh-Bohmer, K., Zhou, X. H., Shirk, S. D., Atri, A., Kukull, W. A., Phelps, C., & Morris, J. C. (2018). Version 3 of the Alzheimer Disease Centers’ Neuropsychological Test Battery in the Uniform Data Set (UDS). Alzheimer Dis Assoc Disord, 32(1), 10–17. 10.1097/WAD.0000000000000223

Whitfield-Gabrieli, S., & Nieto-Castanon, A. (2012). Conn: a functional connectivity toolbox for correlated and anticorrelated brain networks. Brain Connect, 2(3), 125–141. 10.1089/brain.2012.0073

Wolk, D. A., & Detre, J. A. (2012). Arterial spin labeling MRI: an emerging biomarker for Alzheimer’s disease and other neurodegenerative conditions. Curr Opin Neurol, 25(4), 421–428. 10.1097/WCO.0b013e328354ff0a

Wu, J. W., Hussaini, S. A., Bastille, I. M., Rodriguez, G. A., Mrejeru, A., Rilett, K., Sanders, D. W., Cook, C., Fu, H., Boonen, R. A., Herman, M., Nahmani, E., Emrani, S., Figueroa, Y. H., Diamond, M. I., Clelland, C. L., Wray, S., & Duff, K. E. (2016). Neuronal activity enhances tau propagation and tau pathology in vivo. Nat Neurosci, 19(8), 1085–1092. 10.1038/nn.4328

Xi, Q., Zhao, X. H., Wang, P. J., Guo, Q. H., & He, Y. (2013). Abnormal intrinsic brain activity in amnestic mild cognitive impairment revealed by amplitude of low-frequency fluctuation: a resting-state functional magnetic resonance imaging study. Chin Med J (Engl), 126(15), 2912–2917. https://www.ncbi.nlm.nih.gov/pubmed/23924467

Xie, L., Wisse, L. E. M., Pluta, J., de Flores, R., Piskin, V., Manjon, J. V., Wang, H., Das, S. R., Ding, S. L., Wolk, D. A., Yushkevich, P. A., & Alzheimer’s Disease Neuroimaging, I. (2019). Automated segmentation of medial temporal lobe subregions on in vivo T1-weighted MRI in early stages of Alzheimer’s disease. Hum Brain Mapp, 40(12), 3431–3451. 10.1002/hbm.24607

Yan, L., Liu, C. Y., Wong, K. P., Huang, S. C., Mack, W. J., Jann, K., Coppola, G., Ringman, J. M., & Wang, D. J. J. (2018). Regional association of pCASL-MRI with FDG-PET and PiB-PET in people at risk for autosomal dominant Alzheimer’s disease. Neuroimage Clin, 17, 751–760. 10.1016/j.nicl.2017.12.003

Yang, L., Yan, Y., Wang, Y., Hu, X., Lu, J., Chan, P., Yan, T., & Han, Y. (2018). Gradual Disturbances of the Amplitude of Low-Frequency Fluctuations (ALFF) and Fractional ALFF in Alzheimer Spectrum. Front Neurosci, 12, 975. 10.3389/fnins.2018.00975

Yassa, M. A. (2014). Ground zero in Alzheimer’s disease. Nat Neurosci, 17(2), 146–147. 10.1038/nn.3631

Yushkevich, P. A., Munoz Lopez, M., Iniguez de Onzono Martin, M. M., Ittyerah, R., Lim, S., Ravikumar, S., Bedard, M. L., Pickup, S., Liu, W., Wang, J., Hung, L. Y., Lasserve, J., Vergnet, N., Xie, L., Dong, M., Cui, S., McCollum, L., Robinson, J. L., Schuck, T., … Insausti, R. (2021). Three-dimensional mapping of neurofibrillary tangle burden in the human medial temporal lobe. Brain, 144(9), 2784–2797. 10.1093/brain/awab262

Zang, Y. F., He, Y., Zhu, C. Z., Cao, Q. J., Sui, M. Q., Liang, M., Tian, L. X., Jiang, T. Z., & Wang, Y. F. (2007). Altered baseline brain activity in children with ADHD revealed by resting-state functional MRI. Brain Dev, 29(2), 83–91. 10.1016/j.braindev.2006.07.002

Zhang, Q., Wang, Q., He, C., Fan, D., Zhu, Y., Zang, F., Tan, C., Zhang, S., Shu, H., Zhang, Z., Feng, H., Wang, Z., & Xie, C. (2021). Altered Regional Cerebral Blood Flow and Brain Function Across the Alzheimer’s Disease Spectrum: A Potential Biomarker. Front Aging Neurosci, 13, 630382. 10.3389/fnagi.2021.630382

Zhang, Z. H., Fang, X. B., Xi, G. M., Li, W. C., Ling, H. Y., & Qu, P. (2010). Calcitonin gene-related peptide enhances CREB phosphorylation and attenuates tau protein phosphorylation in rat brain during focal cerebral ischemia/reperfusion. Biomed Pharmacother, 64(6), 430–436. 10.1016/j.biopha.2009.06.009

Zhao, Z., Lu, J., Jia, X., Chao, W., Han, Y., Jia, J., & Li, K. (2014). Selective changes of resting-state brain oscillations in aMCI: an fMRI study using ALFF. Biomed Res Int, 2014, 920902. 10.1155/2014/920902

Zhao, Z. L., Fan, F. M., Lu, J., Li, H. J., Jia, L. F., Han, Y., & Li, K. C. (2015). Changes of gray matter volume and amplitude of low-frequency oscillations in amnestic MCI: An integrative multi-modal MRI study. Acta Radiol, 56(5), 614–621. 10.1177/0284185114533329

Zhou, Y., Yu, F., Duong, T. Q., & Alzheimer’s Disease Neuroimaging, I. (2015). White matter lesion load is associated with resting state functional MRI activity and amyloid PET but not FDG in mild cognitive impairment and early Alzheimer’s disease patients. J Magn Reson Imaging, 41(1), 102–109. 10.1002/jmri.24550

Zhuang, L., Liu, X., Xu, X., Yue, C., Shu, H., Bai, F., Yu, H., Shi, Y., & Zhang, Z. (2012). Association of the interleukin 1 beta gene and brain spontaneous activity in amnestic mild cognitive impairment. J Neuroinflammation, 9, 263. 10.1186/1742-2094-9-263

Zuo, X. N., Di Martino, A., Kelly, C., Shehzad, Z. E., Gee, D. G., Klein, D. F., Castellanos, F. X., Biswal, B. B., & Milham, M. P. (2010). The oscillating brain: complex and reliable. Neuroimage, 49(2), 1432–1445. 10.1016/j.neuroimage.2009.09.037

