## Supplementary material for "BOLD Amplitude Correlates of Preclinical Alzheimer’s Disease": Suppl. Fig.

### SUPPLEMENTARY TABLES

**Suppl. Table 1.** ASHS-T1 structural measurements, separated by group.

| | BA36 thickness (mm) | BA35 thickness (mm) | ERC thickness (mm) | aHP volume ($cm^3$) | pHP volume ($cm^3$) | PHC thickness (mm) |
|---|---|---|---|---|---|---|
| ***Group means, adjusted for age and sex*** | | | | | | |
| $M_{A\beta- CU}$ ($N = 131$) | 2.346 | 2.226 | 2.057 | 1.681 | 1.629 | 2.072 |
| $M_{A\beta+ CU}$ ($N = 33$) | 2.363 | 2.264 | 2.093 | 1.751 | 1.605 | 2.052 |
| $M_{A\beta+ MCI}$ ($N = 40$) | 2.218 | 2.056 | 1.878 | 1.456 | 1.354 | 1.980 |
| ***Pairwise comparisons*** | | | | | | |
| Aβ− CU vs. Aβ+ CU (*P*-value) | 0.670 | 0.298 | 0.329 | 0.150 | 0.471 | 0.517 |
| Aβ− CU vs. Aβ+ MCI (*P*-value) | < 0.001 | < 0.001 | < 0.001 | < 0.001 | < 0.001 | 0.001 |
| Aβ+ CU vs. Aβ+ MCI (*P*-value) | 0.002 | < 0.001 | < 0.001 | < 0.001 | < 0.001 | 0.046 |

Abbreviations: aHP = anterior hippocampus; BA36/35 = Brodmann area 36/35; CU = cognitively unimpaired; ERC = entorhinal cortex; MCI = mild cognitive impairment; PHC = parahippocampal cortex; pHP = posterior hippocampus.

**Suppl. Table 2.** Association between MTL tau burden and MTL macrostructural measures in cognitively unimpaired older adults.

| | BA36 | BA35 | ERC | aHP | pHP | PHC |
|---|---|---|---|---|---|---|
| standardized $\beta$ | -0.096 | -0.057 | -0.023 | 0.031 | -0.184 | -0.053 |
| *t*-statistic | -1.088 | -0.621 | -0.264 | 0.390 | -2.291 | -0.587 |
| FWE-corrected *P* | 0.825 | 0.985 | 1.000 | 0.998 | 0.126 | 0.989 |

Macrostructure = thickness for BA36, BA35, ERC, and PHC; macrostructure = volume for aHP and pHP. Abbreviations: aHP = anterior hippocampus, BA36/35 = Brodmann area 36/35, ERC = entorhinal cortex, MTL = medial temporal lobe, PHC = parahippocampal cortex, pHP = posterior hippocampus.

**Supp. Table 3.** Association between MTL tau and MTL ALFF in individuals with less than 1.5-year interval between fMRI and tau-PET scans.

| | BA36 | BA35 | ERC | aHP | pHP | PHC |
|---|---|---|---|---|---|---|
| **Baseline model (*N* = 122, *df* = 113)** | | | | | | |
| standardized $\beta$ | -0.119 | -0.156 | -0.224 | -0.138 | -0.122 | -0.131 |
| *t*-statistic | -1.443 | -1.892 | -2.857 | -1.697 | -1.502 | -1.630 |
| FWE-corrected *P* | 0.347 | 0.161 | 0.019 | 0.230 | 0.316 | 0.257 |
| **Controlling for structural measures (*N* = 122, *df* = 111/112)** | | | | | | |
| standardized $\beta$ | -0.125 | -0.176 | -0.242 | -0.133 | -0.159 | -0.107 |
| *t*-statistic | -1.519 | -2.188 | -3.549 | -2.264 | -2.216 | -1.569 |
| FWE-corrected *P* | 0.380 | 0.107 | 0.003 | 0.151 | 0.098 | 0.247 |
| **Controlling for structural measures and amyloid status (*N* = 122, *df* = 109/110)** | | | | | | |
| standardized $\beta$ | -0.030 | -0.125 | -0.240 | -0.154 | -0.171 | -0.082 |
| *t*-statistic | -0.304 | -1.287 | -2.903 | -1.901 | -2.006 | -0.869 |
| FWE-corrected *P* | 0.999 | 0.527 | 0.020 | 0.195 | 0.159 | 0.822 |

Abbreviations: ALFF = amplitude of low-frequency fluctuations, aHP = anterior hippocampus, BA36/35 = Brodmann area 36/35, ERC = entorhinal cortex, MTL = medial temporal lobe, PHC = parahippocampal cortex, pHP = posterior hippocampus.

**Suppl. Table 4.** Association between MTL tau and MTL ALFF in cognitively unimpaired older adults with less than 1.5-year interval between fMRI and tau-PET scans.

| | **BA36** | **BA35** | **ERC** | **aHP** | **pHP** | **PHC** |
|---|---|---|---|---|---|---|
| **All CU: Baseline model (*N* = 114, *df* = 105)** | | | | | | |
| standardized $\beta$ | -0.136 | -0.147 | -0.236 | -0.154 | -0.105 | -0.121 |
| *t*-statistic | -1.593 | -1.727 | -2.973 | -1.855 | -1.239 | -1.453 |
| FWE-corrected *P* | 0.283 | 0.224 | 0.013 | 0.178 | 0.475 | 0.352 |
| **All CU: Controlling for structural measures (*N* = 114, *df* = 103/104)** | | | | | | |
| standardized $\beta$ | -0.142 | -0.153 | -0.232 | -0.124 | -0.113 | -0.132 |
| *t*-statistic | -1.673 | -1.839 | -3.426 | -1.855 | -1.610 | -1.667 |
| FWE-corrected *P* | 0.298 | 0.221 | 0.004 | 0.211 | 0.326 | 0.300 |
| **All CU: Controlling for structural measures and amyloid status (*N* = 114, *df* = 101/102)** | | | | | | |
| standardized $\beta$ | -0.074 | -0.111 | -0.232 | -0.133 | -0.125 | -0.096 |
| *t*-statistic | -0.800 | -1.211 | -3.080 | -1.798 | -1.602 | -1.098 |
| FWE-corrected *P* | 0.858 | 0.581 | 0.012 | 0.234 | 0.327 | 0.660 |

Abbreviations: ALFF = amplitude of low-frequency fluctuations, aHP = anterior hippocampus, BA36/35 = Brodmann area 36/35, ERC = entorhinal cortex, MTL = medial temporal lobe, PHC = parahippocampal cortex, pHP = posterior hippocampus.

**Suppl. Table 5.** Framewise displacement (FD) for raw and filtered realignment parameters, separated by group.

| | **Aβ− CU (SD)** | **Aβ+ CU (SD)** | **Aβ+ MCI (SD)** | **Statistical Differences** |
|---|---|---|---|---|
| Raw mean FD (mm) | 0.2895 (0.1657) | 0.2473 (0.1133) | 0.2083 (0.0821) | Aβ− CU > Aβ+ MCI *** |
| Raw max FD (mm) | 1.2085 (1.0001) | 1.1242 (0.6771) | 1.1197 (1.0626) | none |
| Filtered mean FD (mm) | 0.0431 (0.0219) | 0.0422 (0.0212) | 0.0350 (0.0216) | none |
| Filtered max FD (mm) | 0.1937 (0.1497) | 0.2036 (0.1380) | 0.1979 (0.1908) | none |

***** *P* < 0.001**

**Suppl. Table 6.** MTL voxelwise temporal Signal-to-Noise Ratio (tSNR) for minimally processed* fMRI datasets, separated by group.

| | BA36 tSNR (SD) | BA35 tSNR (SD) | ERC tSNR (SD) | aHP tSNR (SD) | pHP tSNR (SD) | PHC tSNR (SD) |
|---|---|---|---|---|---|---|
| ***Group means, adjusted for age and sex*** | | | | | | |
| Aβ− CU | 12.407 (2.466) | 10.569 (2.251) | 8.643 (2.072) | 15.928 (2.749) | 19.944 (2.275) | 18.418 (2.973) |
| Aβ+ CU | 12.978 (1.917) | 10.863 (2.379) | 8.469 (1.830) | 16.033 (3.128) | 20.390 (2.066) | 19.584 (2.339) |
| Aβ+ MCI | 12.373 (2.055) | 10.282 (1.963) | 8.500 (1.770) | 15.810 (2.625) | 20.602 (2.095) | 19.062 (2.018) |
| Statistical differences | none | none | none | none | none | none |

*Minimal processing consisted of field map-based distortion correction, motion correction using realignment, and structural-functional rigid-body registration in native space. Abbreviations: aHP = anterior hippocampus, BA36/35 = Brodmann area 36/35, ERC = entorhinal cortex, MTL = medial temporal lobe, PHC = parahippocampal cortex, pHP = posterior hippocampus.

### SUPPLEMENTARY FIGURES

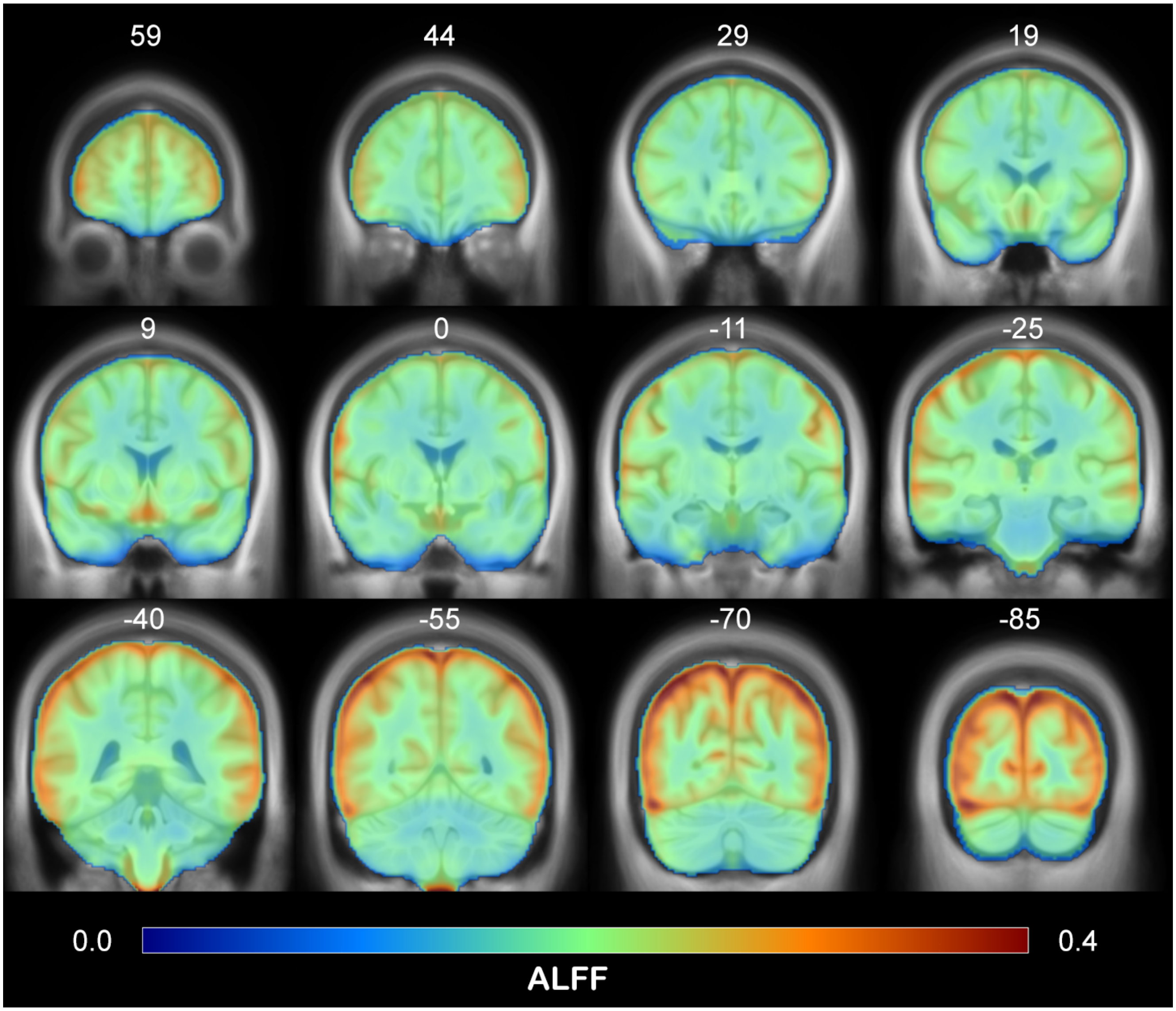


**Suppl. Figure 1.** Raw amplitude of low-frequency fluctuations (ALFF) map, averaged across participants.

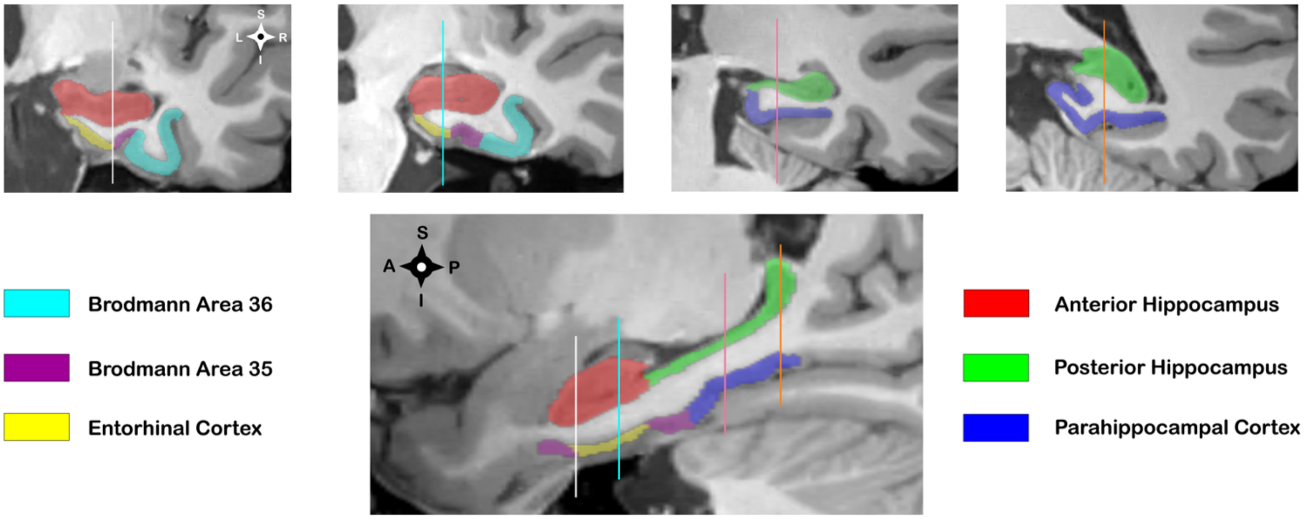


**Suppl. Figure 2.** Example ASHS-T1 segmentation from a single participant.

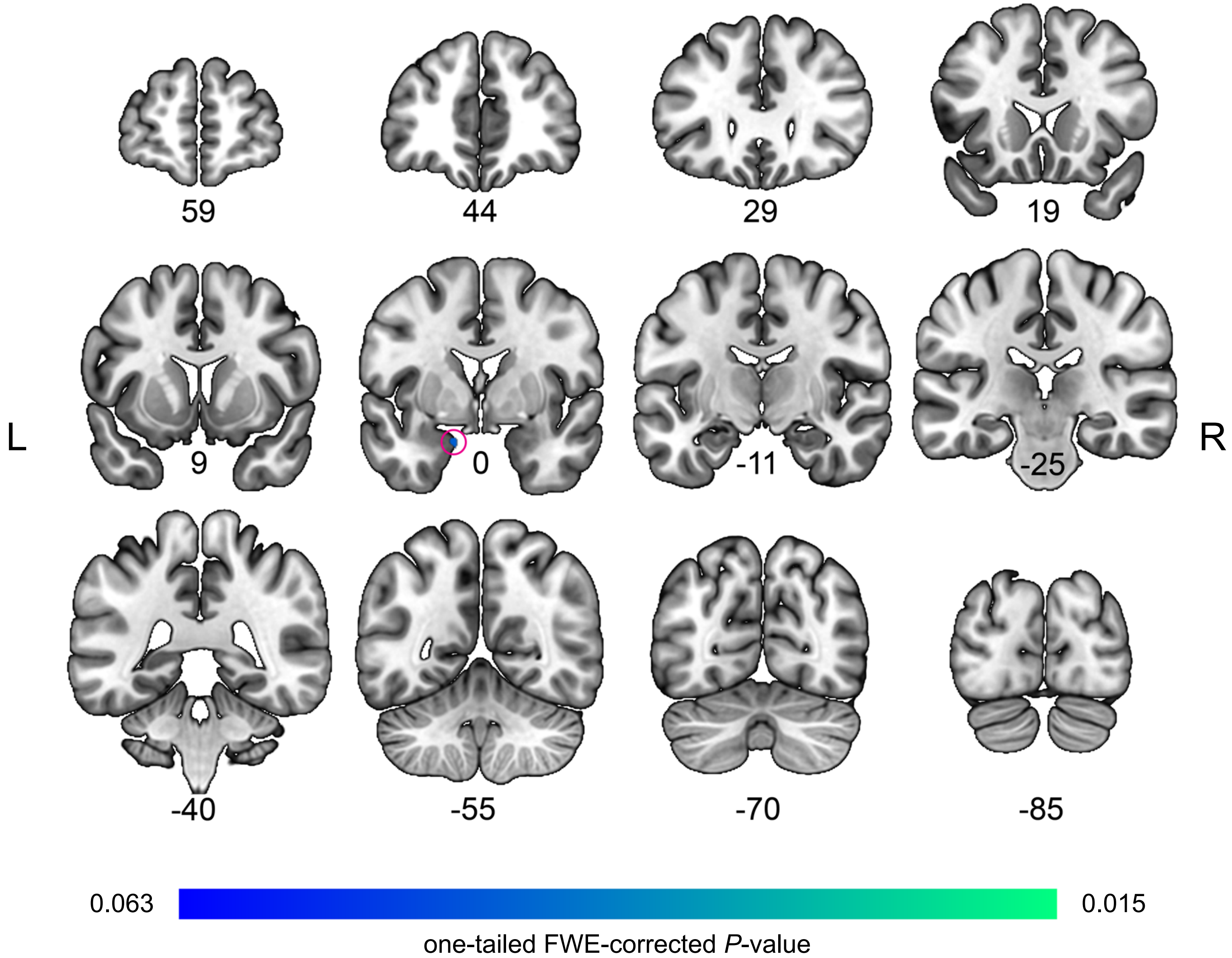


**Suppl. Figure 3.** Voxelwise tests of association between tau burden in the MTL and cortical ALFF in cognitively unimpaired amyloid-positive and amyloid-negative older adults. Voxelwise GLMs were performed with age, sex, and head movement metrics as nuisance covariates. The color map corresponds to one-tailed FWE-corrected TFCE *P*-values thresholded at $\alpha = 0.050$. Abbreviations: ALFF = amplitude of low-frequency fluctuations; TFCE = threshold-free cluster enhancement.

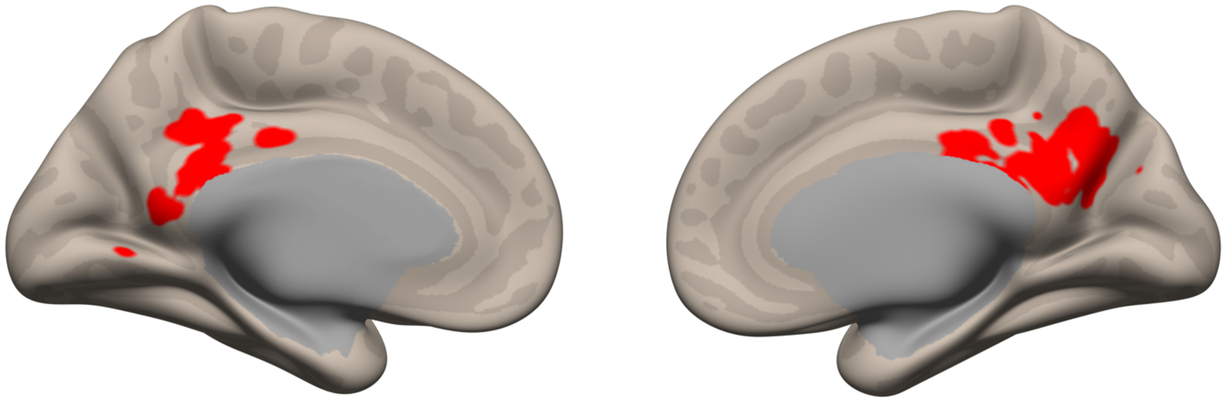

**Suppl. Figure 4.** Medial parietal ROI that was used in the mediation analysis investigating the relationships among the amyloid group (i.e., Aβ-positive or Aβ-negative), tau, and ALFF. This figure is a complement to Fig. 5 in the main text.
